# Primary resistance to ICI-based regimens is associated with early longitudinal changes in the fecal microbiome and loss of microbial stability

**DOI:** 10.1101/2025.10.01.25336378

**Authors:** Yujie Zhao, Jarushka Naidoo, Michael Conroy, Jacqueline T Ferri, Joell J Gills, Krista Y Chen, James R White, Sara Glass, William O Assan, Kimberly Peloza, Megan D Schollenberger, William H Sharfman, Kristen A Marrone, Patrick Forde, Julie Brahmer, Valsalmo Anagnostou, Drew M Pardoll, Joseph Murray, Evan J Lipson, Cynthia L Sears, Fyza Y Shaikh

## Abstract

While immune checkpoint inhibitors (ICI)-based regimens and chemo-immunotherapy combinations (chemo-ICI) are now first-line therapy for patients with metastatic non-small cell lung cancer (NSCLC), the number of patients who experience a sustained response to treatment remains limited. The majority of patients will not benefit from initial treatment (primary resistance) or develop progressive disease after an initial period of response (acquired resistance). Microbiome-based biomarkers offer an opportunity to identify patients who may have a poor response to ICI-based regimens using non-invasive methods, and potentially, then the microbiota may be amenable to therapy-enhancing alteration. However, a deep understanding of the longitudinal dynamics of gut microbial features in patients treated with ICIs in NSCLC, a feature of likely importance for microbiome-based therapeutics, remains limited. In this study, we show that patients with NSCLC who experience primary resistance to an ICI-based regimen show a loss of intra-individual microbiome stability in the first 4 months during treatment with an ICI-based regimen, independent of antibiotic exposure. Loss of microbiome stability was validated in a second tumor type in patients with melanoma. To identify key microbial species associated with progression, recursive feature elimination with random forest classifiers was used to identify temporally associated microbial species associated with disease progression. An index of these progression-associated species was able to predict clinical outcomes based on pre-treatment fecal samples, with further validation on an independent cohort. Together, our data show that microbial instability may be an early indicator of ICI-resistance in patients with NSCLC and melanoma, with the potential to be developed into biomarkers of primary resistance to ICI-based regimens.

## INTRODUCTION

Immune checkpoint inhibitors (ICIs) have improved treatment outcomes for patients with non-small cell lung cancer (NSCLC). Although some patients with NSCLC will have durable responses to ICI-based regimens, the majority of patients do not benefit from initial treatment (primary resistance) or develop progressive disease after an initial period of response (acquired resistance)^1^. In newly diagnosed NSCLC, several approved regimens with ICI monotherapy, ICI dual therapy, or chemo-ICI have shown benefit in both progression-free survival (PFS) and overall survival (OS). However, only 15-20% of these patients will experience long-term durable benefit from therapy^2–7^. Several genetic alterations have been linked with acquired resistance, such as STK11 mutation, beta-2-microglobulin and human leukocyte antigen class I loss^8,9^. However, there are currently no targeted therapies to address either primary or acquired resistance, and biomarkers to predict resistance and guide treatment remain limited to unmodifiable factors.

The gut microbiome is putatively amenable to therapy-enhancing alteration through use of prebiotics (i.e. fiber supplements), tailored live bacterial microbiota therapy (e.g., SER-109, now Vowst), or other broad changes in diet (i.e. plant-based vs meat-based)^10–12^. Multiple studies suggest that the gut microbiome may be utilized as a biomarker or may even modulate the anti-tumor immune response during ICI treatment. Modification of the microbiome using fecal microbiota transfer has shown promise in overcoming immunotherapy resistance in melanoma, and higher fiber intake has been associated with improved overall survival^13–15^. These data indicate a critical role for a modifiable gut microbiome during ICI-treatment.

For ICI therapy in NSCLC, the specific bacteria or bacterial communities both putatively helpful, or perhaps more importantly harmful, in ICI responses remain inconsistent across study populations and tumor types^16–25^. Most prior studies have used a cross-sectional approach to analyze fecal samples collected at the time of treatment initiation, and thus, are limited in understanding dynamic shifts during ICI treatment. Two recent reports in melanoma described longitudinal microbiome associations. Higher abundances of some bacterial species were confirmed in durable responses based on longitudinal trajectory, including *Faecalibacterium prausnitzii* and *Bacteroides thetaiotaomicron*, while others were found to increase in those who had disease progression within a year, e.g. *Parabacteroides distasonis and Ruminococcus torques* (n=175 patients)^26^. The stability of the microbiome has also been described in melanoma patients, specifically in patients with durable responses (n=23 patients)^27^. Data for longitudinal dynamics and stability in NSCLC remain limited, with one study (n=37) finding no change in gut microbiome composition from pre-treatment compared to time of progression^18^.

Herein, we present data from a NSCLC cohort (n=101 patients) indicating early loss of microbiome stability in the first 4 months as a potential indicator of poor response to treatment in patients receiving ICI-based regimens, independent of antibiotic exposure at treatment initiation. We utilized fecal samples at the time of progression to identify progression-associated bacterial species, and further, correlated an index of these species with progression-free survival (PFS) and overall survival (OS) in an independent NSCLC cohort. Additionally, we validate our findings for microbiome stability in a second tumor cohort of patients with melanoma (n=28). These findings support the importance of early longitudinal microbiome shifts as a potential tool for biomarker development and as a therapeutic target for microbiome-based interventions.

## RESULTS

### NSCLC cohort characteristics

A total of 101 patients with 374 total fecal samples were evaluated for microbiome features based on clinical response to an ICI-based regimen (Table 1, Table S1). Patients were divided into 4 groups based on PFS. Response groups consisted of primary resistance to an ICI-based regimen (PR, n=47, PFS < 6 months), acquired resistance after 6 months of partial response or stable disease (AR6, n=19, PFS > 6 months but < 12 months), acquired resistance after 12 months of partial response or stable disease (AR12, n=21, PFS > 12 months with eventual progression), and durable clinical benefit or response (DR, n=14, PFS >12 months without progression at time data was censored). The majority of patients across tumor response groups had metastatic disease at the time of treatment initiation (74-94%) and were current or former smokers (74-100%). Patients in each group were most often treated with ICI as a single agent (42-71%); and the rate of immune-related adverse events (irAEs) ranged widely (15-68%). A longitudinal distribution of samples is shown by timepoints in 60-day intervals (Fig S1A), as well as total number of samples per patient (Fig S1B) and distribution of samples relative to start of treatment for each patient (Fig S1C).

**Table 1.** Non-small cell lung cancer cohort characteristics by tumor response. Abbreviations. irAE, immune related adverse event.

| Characteristic | Overall<br>N = 101 | Primary<br>Resistance (PR)<br>N = 47<sup>†</sup> | Acquired Resistance<br>after 6 months (AR6)<br>N = 19<sup>†</sup> | Acquired Resistance<br>after 12 months (AR12)<br>N = 21<sup>†</sup> | Durable<br>Response (DR)<br>N = 14<sup>†</sup> |
| --- | --- | --- | --- | --- | --- |
| Age [yrs] (SD) | 65 (9) | 65 (10) | 66 (9) | 67 (9) | 62 (8) |
| Sex (%) |  |  |  |  |  |
| Female | 55 (54%) | 27 (57%) | 11 (58%) | 10 (48%) | 7 (50%) |
| Male | 46m (46%) | 20 (43%) | 8 (42%) | 11 (52%) | 7 (50%) |
| Race (%) |  |  |  |  |  |
| Black | 16 (16%) | 8 (17%) | 3 (16%) | 2 (9.5%) | 3 (21%) |
| White | 79 (78%) | 36 (77%) | 15 (79%) | 18 (86%) | 10 (71%) |
| Other | 6 (6%) | 3 (6.4%) | 1 (5.3%) | 1 (4.8%) | 1 (7.1%) |
| Stage (%) |  |  |  |  |  |
| Advanced (III/IIIA/IIIB) | 15 (15%) | 3 (6.4%) | 5 (26%) | 5 (24%) | 2 (14%) |
| Metastatic (IV) | 86 (85%) | 44 (94%) | 14 (74%) | 16 (76%) | 12 (86%) |
| Smoking (%) |  |  |  |  |  |
| Current/Former | 82 (81%) | 35 (74%) | 17 (89%) | 16 (76%) | 14 (100%) |
| Never | 19 (19%) | 12 (26%) | 2 (11%) | 5 (24%) | 0 (0%) |
| Histology (%) |  |  |  |  |  |
| Adenocarcinoma | 72 (71%) | 35 (74%) | 9 (47%) | 17 (81%) | 11 (79%) |
| Squamous | 23 (23%) | 9 (19%) | 8 (42%) | 4 (19%) | 2 (14%) |
| Other | 6 (6%) | 3 (6.4%) | 2 (11%) | 0 (0%) | 1 (7.1%) |
| ECOG (%) |  |  |  |  |  |
| 0-1 | 90 (89%) | 41 (87%) | 16 (84%) | 19 (90%) | 14 (100%) |
| 2+ | 11 (11%) | 6 (13%) | 3 (16%) | 2 (9.5%) | 0 (0%) |
| Treatment |  |  |  |  |  |
| ICI single agent | 57 (56%) | 25 (53%) | 8 (42%) | 15 (71%) | 9 (64%) |
| ICI combination | 6 (6%) | 2 (4.3%) | 3 (16%) | 0 (0%) | 1 (7.1%) |
| ICI + chemotherapy | 28 (28%) | 14 (30%) | 6 (32%) | 5 (24%) | 3 (21%) |
| ICI + other agent<sup>2</sup> | 10 (10%) | 6 (13%) | 2 (11%) | 1 (4.8%) | 1 (7.1%) |
| Treatment Line |  |  |  |  |  |
| 1 | 40 (39.6%) | 16 (36%) | 7 (41%) | 10 (53%) | 7 (54%) |
| 2 | 35 (34.7%) | 15 (33%) | 6 (35%) | 9 (47%) | 5 (38%) |
| 3+ | 19 (18.8%) | 14 (31%) | 4 (24%) | 0 (0%) | 1 (7.7%) |
| (Missing)<sup>3</sup> | 7 (6.9%) | 2 | 2 | 2 | 1 |
| Treatment History |  |  |  |  |  |
| Prior Chemotherapy | 60 (59%) | 31 (66%) | 12 (63%) | 11 (52%) | 6 (43%) |
| Prior ICI | 14 (14%) | 8 (17%) | 3 (16%) | 1 (4.8%) | 2 (14%) |
| Outcomes |  |  |  |  |  |
| Progression | 85 (84%) | 45 (96%) | 19 (100%) | 21 (100%) | 0 (0%) |
| Death | 70 (69%) | 42 (89%) | 16 (84%) | 11 (52%) | 1 (7.1%) |
| irAE | 40 (40%) | 7 (15%) | 13 (68%) | 11 (52%) | 9 (64%) |
| Tumor Mutation Burden<br>(TMB) |  |  |  |  |  |
| High | 13 (12.9%) | 4 (14%) | 2 (20%) | 2 (15%) | 5 (45%) |
| Intermediate | 22 (21.8%) | 8 (28%) | 6 (60%) | 4 (31%) | 4 (36%) |
| Low | 28 (27.7%) | 17 (59%) | 2 (20%) | 7 (54%) | 2 (18%) |
| (Missing) | 38 (37.6%) | 18 | 9 | 8 | 3 |
| Tumor Proportion Score<br>(TPS) |  |  |  |  |  |
| TPS <1 | 26 (25.7%) | 12 (35%) | 3 (25%) | 6 (32%) | 5 (36%) |
| TPS 1-49 | 17 (16.8%) | 8 (24%) | 3 (25%) | 5 (26%) | 1 (7.1%) |
| TPS 50-100 | 36 (35.6%) | 14 (41%) | 6 (50%) | 8 (42%) | 8 (57%) |
| (Missing) | 22 (21.8%) | 13 | 7 | 2 | 0 |

<sup>†</sup> Mean (SD); n (%)<sup>2</sup> Monalizumab, Olecumab, STAT3 inhibitor, azacitidine+entinostat, ceralasertib, IDO inhibitor, linrodostat<sup>3</sup> ICI as consolidation therapy

Clinical characteristics associated with PFS and OS in this cohort were assessed using a Cox Proportional Hazards model (Table S2 and S3, respectively). The results indicate that later lines of treatment were associated with shorter PFS; while development of any-grade irAEs during treatment and smoking exposure were associated with longer PFS (Fig 1A, Table S2). Of these, development of irAEs remained significant relative for OS, while treatment line showed a similar trend (p = 0.11, Table S3). Tumor mutational burden (TMB) showed a trend toward longer PFS and OS (p = 0.08 and p = 0.18, respectively, Tables S2-3). Overall survival stratified by tumor response groups (as described in Table 1) was also significant (Fig 1B).

**Figure 1.**
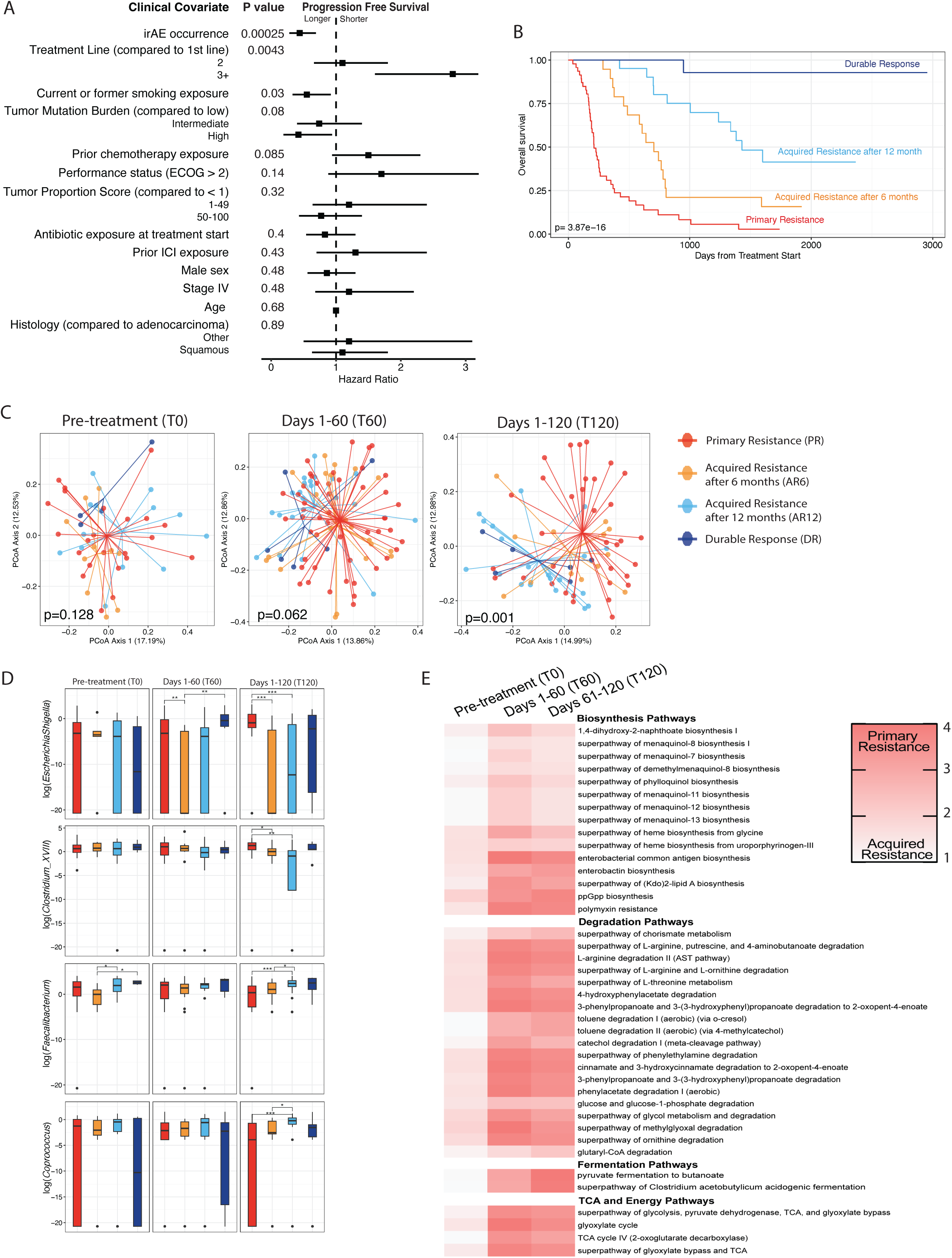
The fecal microbiome shifts in patients with primary resistance to ICI therapy by 120 days after start of treatment. ***A***. Clinical co-variates identified by univariate cox regression relative to progression free survival. Square represents the hazard ratio and the horizontal bars represent the 95% confidence interval. Statistics: likelihood ratio test, with additional statistics in Table S2. ***B***. Kaplan-Meier curve for overall survival probability by tumor response group. Statistics: Log-Rank test. ***C***. Principal Coordinate Analysis (PCoA) of fecal samples representing Bray-Curtis dissimilarity metric across pre-treatment samples (T0, left, n= 50 samples for 50 individuals), samples collected between day 1 to day 60 (T60, middle, n=98 samples for 70 individuals, middle), samples collected between day 61 to day 120 (right, T120, n=73 samples for 52 individuals). Color indicates response group. Statistics: Permutational Multivariate Analysis of Variance (PERMANOVA). ***D***. The abundance of individual genera, measured by 16S rRNA amplicon sequencing for fecal samples collected by response group (color) at different treatment timepoints: pre-treatment (T0, left); days 1-60 (T60, middle); days 61-120 (T120, right). Boxplot represents median, 25% to 75% interquartile range (bottom and top edge), and 95% confidence intervals (whiskers). Statistics: Mann-Whitney test. ***E***. Heatmap showing longitudinal fold change (log_2_) for functional pathways enriched in primary resistance. Pathways selected for heatmap were significant using Mann Whitney test at both T60 and T120 between primary resistance (PR) and acquired resistance at 6 months group (AR6). Data corresponds to Table S5.

### Longitudinal microbiome shifts in patients with primary resistance

To assess longitudinal microbiome shifts by tumor response group, all fecal samples were classified into 60-day intervals based on date of sample collection relative to treatment start for each patient. In the primary resistance group, fecal samples were limited after disease progression (Fig S1A), thus direct comparisons which included the primary resistance group were focused on the first 180 days of treatment. Initial analysis of the overall microbiome composition (Bray-Curtis) between tumor response groups shows that the fecal microbiome in patients with primary resistance shifts away from other groups in the first 120 days on treatment (Fig 1C). This shift begins at days 1-60 (Fig 1C; T60, p = 0.062) and shifts further by days 61-120 (Fig 1C, T120, p = 0.01). Of note, the difference in microbiome profiles is not evident at pre-treatment (Fig 1C, T0, p = 0.128).

To identify the longitudinal genera driving this change in overall composition, we performed genus-level pairwise comparisons at day 61-120 (T120) with correction for false discovery rate (FDR, Table S4). Four genera were identified as significant across tumor response groups. The relative abundances of *Escherichia-Shigella* and *Clostridium_XVIII* were enriched in primary resistance, while *Faecalibacterium* and *Coprococcus* were enriched in acquired resistance (Fig 1D, S2A). As *Escherichia* 16S rRNA amplicons can be ambiguous, the sequencing data was also validated with qPCR (Fig S2B); and qPCR results correlated with 16S rRNA amplicon sequencing data (Fig S2C). Notably, these genera were not consistently different between tumor response groups at pre-treatment or earlier timepoints (Fig 1D, left and middle panels), and alpha diversity did not show significant changes across timepoints (Fig S2D-E). A similar analysis was also performed for predicted functional pathways using PICRUSt2^28^. Pathways that were not significant at pre-treatment timepoints became enriched in the primary resistance group at subsequent timepoints by day 120 (Fig 1E, Table S5). These pathways have a wide range of function, including vitamin K synthesis, degradation of amino acids and aromatic compounds, and bacterial fermentation.

### Antibiotic exposure at the time of treatment initiation

As antibiotics have been implicated in gut microbiome-mediated effects on progression and survival^29,30^, we explored the microbiome composition stratified by antibiotic exposure and among each response group. When analyzing all samples collectively, antibiotic exposure significantly altered microbiome composition at all timepoints (Fig. S3A), indicating an overall effect on the microbiome consistent with expected bacterial compositional changes as a result of antibiotic treatment. However, the bacterial composition shifts seen between tumor response groups remained significant at days 1-60 (T60) and 61-120 (T120) independent of antibiotic exposure (Fig S3B-C). To better define risk for each response group, we also performed Cox regression analysis using both PFS and OS by treatment response group (Fig S4). While antibiotic exposure did not appear to influence PFS, the hazard ratio relative to OS was notable for association with both antibiotic exposure at treatment initiation (PR; primary resistance group) and before progression (AR; acquired resistance group) are consistent with a subtle, but likely pervasive impact, of antibiotic exposure on the duration of OS (Fig S4D). Collectively, these data indicate that microbial shifts occurring at treatment initiation are both antibiotic-independent and -dependent. Antibiotic-dependent microbial changes appear to influence OS in both primary and acquired resistance, consistent with prior reports^17,29–32^.

### Intra-individual microbiome stability is lost in primary resistance

To assess the fecal microbiome stability between tumor response groups, we utilized three analytic tools. First, we applied the intra-class correlation (ICC) analysis to all fecal samples which assesses the similarity across all samples for an individual patient. A higher ICC score indicates the microbiome composition of samples from one patient are more similar to each other, while a lower score indicates more variability between samples from the same patient. Notably, the ICC score was lower for the primary resistance group compared to both acquired resistance groups (AR6, AR12), indicating fecal samples from the same patient had greater variability (Fig 2A, p = 0.045 and p=0.028, respectively). The durable response group was not significantly different from any of the other groups, possibly due to smaller sample size (see figure legend) and/or longer overall collection period as durable response patients remained on ICI-based regimens for a longer duration (Fig S1C). Second, we examined compositional shifts within individual patients from pre-treatment to on therapy timepoints using Bray-Curtis dissimilarity. The fecal microbiome of patients with primary resistance shifted away from pre-treatment composition at both days 1-60 and 61-120 (Fig 2B-C), while this shift did not occur in patients with acquired resistance (Fig 2D-E).

**Figure 2.**
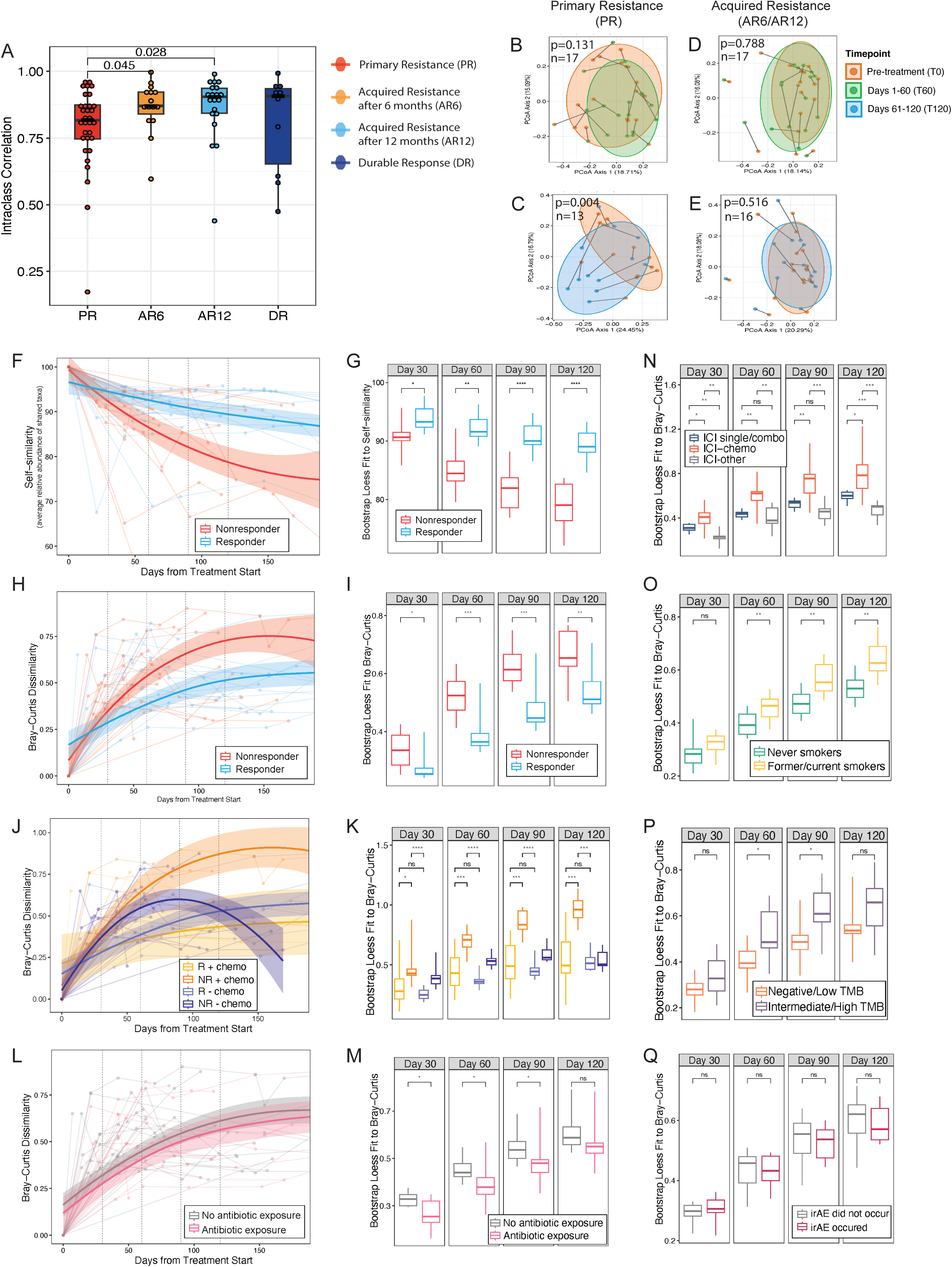
Fecal samples in NSCLC primary resistance show loss of intra-individual taxonomic stability from baseline. ***A***. Intraclass correlation (ICC) for similarity of longitudinal fecal samples within each patient (n=77 patients). ***B-E***. Principal Coordinate Analysis (PCoA) of microbiome composition using Bray-Curtis distance metric. Microbiome composition shifts in individual patients were directly compared from pre-treatment fecal samples (T0) to on-therapy timepoints (either T60 or T120). B-C show the shifts for primary resistance group while D-E show acquired resistance (AR6 and AR12). Statistics performed using Permutational Multivariate Analysis of Variance (PERMANOVA) with the correction for individual subjects. ***F-M.*** Microbiome distance metrics are shown using a locally estimated scatterplot smoothing (LOESS) model for fecal samples sorted by tumor response at 6 months into nonresponders (primary resistance) and responders (acquired resistance and durable response). Individual samples are shown with lines connecting each patient. Data from LOESS curves was quantified at days 30, 60, 90, and 120 using bootstrapping. Distance metrics include self similarity (F-G); Bray-Curtis dissimilarity (H-I); Bray-Curtis dissimilarity in responders and nonresponders stratified by chemotherapy exposure (J-K), and Bray-Curtis dissimilarity stratified by antibiotic exposure +/- 42 days of treatment start (L-M). **N-Q**. Additional LOESS quantification of Bray-Curtis dissimilarity using bootstrapping for treatment regimens (N), smoking exposure (O), tumor mutational burden (P), and immune-related adverse events (O). ***Abbreviations***. PR, primary resistance; AR6, acquired resistance after 6 months of partial response or stable disease, AR12, acquired resistance after 12 months of partial response or stable disease; DR, durable response; TMB, tumor mutation burden; irAE, immune-related adverse event.

Third, since our longitudinal samples were not collected at pre-determined timepoints, we used several metrics to evaluate the overall microbiome composition and fit the resulting data into a locally estimated scatterplot smoothing (LOESS) model. Bray-Curtis and UniFrac were used to evaluate dissimilarity, with higher values indicates larger shifts from pre-treatment composition; while self-similarity (see methods) was used to evaluate shared species, with lower values indicating a larger shift from pre-treatment composition. The resulting LOESS curves were quantified at days 30, 60, 90, and 120 using bootstrapping. To focus on shifts in primary resistance in the first 180 days, patients were sorted based on response at 6 months, with primary resistance group classified as nonresponders (PFS < 6 months) while acquired resistance and durable response were classified into responders (PFS > 6 months).

The data show that the fecal microbiome of nonresponders was less stable and rapidly shifted away from their own pre-treatment profile using self-similarity (Fig 2F-G) and Bray-Curtis dissimilarity (Fig 2H-I, S5). These longitudinal shifts were not driven by antibiotic exposure, as antibiotic exposure alone did not alter dissimilarity (Fig 2L-M). Since these tumor response groups include both ICI monotherapy and chemo-ICI, we further stratified responders and nonresponders by ICI therapies alone or ICI plus chemotherapy exposure (Fig J-K). Early microbiome instability after therapy initiation was observed in nonresponders regardless of whether treated with ICI therapy alone or ICI plus chemotherapy. However, the association of nonresponse with microbiome instability was accentuated in those treated with ICIs plus chemotherapy. Together, these data suggest that nonresponse is a strong driver of microbiome instability with additional potential impact by treatment regimen. We further examined other clinical characteristics. Combination chemo-ICI, smoking status, and TMB, all correlated with higher Bray-Curtis dissimilarity (Fig 2N-P), while irAEs did not (Fig 2Q). Additional results for each co-variate, as well as results for unweighted and weighted UniFrac, are shown in the supplementary data (Fig S5-8). Fig S5 and S6 reveal that, regardless of metric, the microbiome in patients with primary resistance displays greater instability or dissimilarity in composition early during therapy. Fig S7 emphasizes the limited impact of antibiotic exposure similar to Fig S3 and S4. Fig S8 displays self-similarity and unweighted and weighted UniFrac data for smoking, TMB and treatment lines. For individual patients, the longitudinal genus-level taxonomic shifts are also shown in Figure S9.

### Microbiome stability in melanoma

To further validate our longitudinal and microbiome stability findings, we evaluated a cohort of patients with melanoma as a second tumor type. We identified 28 patients (93 total samples) with advanced/unresectable or metastatic melanoma who had fecal samples at pre-treatment and additional longitudinal timepoints (Table 2). Clinical characteristics were similar between patients with response or non-response at 6 months with most patients treated on ICI monotherapy (65-75%). Notably, irAEs were present in 100% of the responders but only in 55% of nonresponders (p = 0.029). We applied both self-similarity (Fig 3A-B) and Bray-Curtis dissimilarity (Fig 3C-D) to this dataset and fit the data into a LOESS model with quantification at early timepoints by bootstrapping. Our data demonstrate that melanoma nonresponders show an early loss of microbiome stability, similar to the results shown for NSCLC (Fig 2), and our data are also consistent with a prior longitudinal study in melanoma^27^. As with our NSCLC data, we did not see shifts in alpha diversity longitudinally between responders and nonresponders (Fig 3E-F). These data are consistent with our prior work re-analyzing raw data from multiple published melanoma cohorts^16^, as well as other reports have finding conflicting results for alpha diversity in both melanoma and NSCLC^16,18,19,22,33^. Further stratification by clinical co-variates indicates the context of stability may be specific to each tumor type. Smoking status (Fig 3G) was not associated with microbiome instability while development of irAEs (Fig 3H) was associated, in contrast to the NSCLC cohort. We also assessed first-line treatment (Fig 3I, associated with instability) and combination ICI therapy without chemotherapy (Fig3J, not associated with instability).

**Figure 3.**
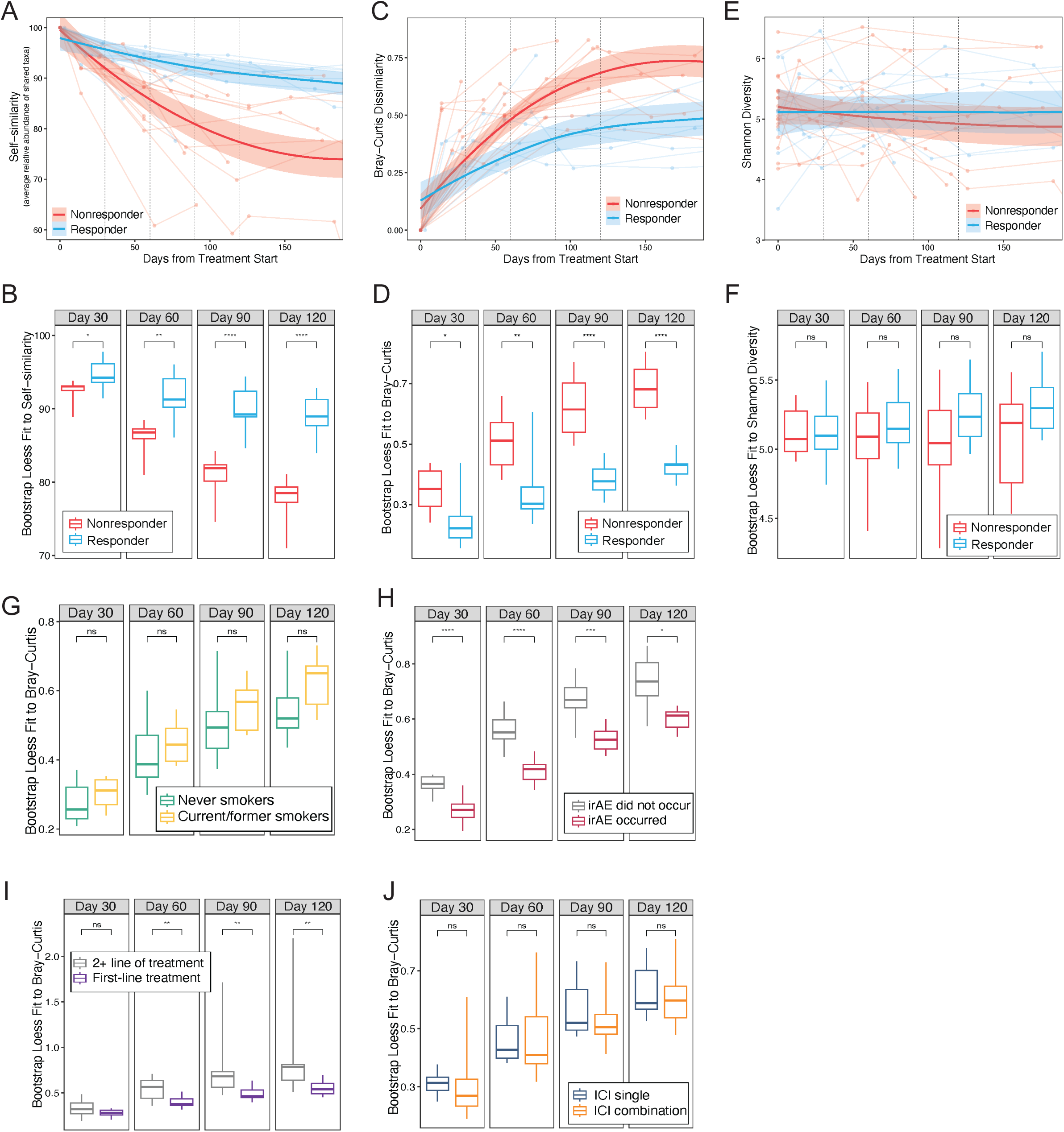
Loss of stability correlates with nonresponse in melanoma, independent of alpha diversity. ***A-D***. Microbiome distance metrics are shown using a locally estimated scatterplot smoothing (LOESS) model for fecal samples sorted by tumor response at 6 months into nonresponders and responders. Individual samples are shown with lines connecting each patient. Data from LOESS curves was quantified at days 30, 60, 90, and 120 using bootstrapping. Distance metrics: self similarity (A-B) and Bray-Curtis dissimilarity (C-D). ***E-F***. LOESS model for alpha diversity is shown stratified by response group. ***G-J***. Quantification of Bray-Curtis dissimilarity using LOESS and bootstrap for clinical co-variates: smoking status (G), and immune-related adverse events (irAE, H), first-line treatment (I), and combination immune checkpoint inhibitor (ICI) regimens without chemotherapy (J)

**Table 2.** Melanoma cohort characteristics by tumor response at 6 months. Abbreviations. irAE, immune-related adverse events.

| Characteristic | Overall<br>N = 28 | Responder<br>N = 8<sup>1</sup> | Nonresponder<br>N = 20<sup>1</sup> | p-value<sup>2</sup> |
| --- | --- | --- | --- | --- |
| Age [yrs] (SD) | 67 (15) | 70 (14) | 66 (16) | 0.6 |
| Sex (%) |  |  |  | 0.4 |
| Female | 12 (43%) | 2 (25%) | 10 (50%) |  |
| Male | 16 (57%) | 6 (75%) | 10 (50%) |  |
| Race (%) |  |  |  | >0.9 |
| White | 27 (96%) | 8 (100%) | 19 (95%) |  |
| Other | 1 (4%) | 0 (0%) | 1 (5.0%) |  |
| Stage (%) |  |  |  | 0.5 |
| Advanced (IIIC) | 3 (11%) | 0 (0%) | 3 (15%) |  |
| Metastatic (IV) | 25 (89%) | 8 (100%) | 17 (85%) |  |
| Smoking (%) |  |  |  | >0.9 |
| Current/Former | 15 (54%) | 4 (50%) | 11 (55%) |  |
| Never | 13 (46%) | 4 (50%) | 9 (45%) |  |
| Treatment (%) |  |  |  | >0.9 |
| ICI single agent | 19 (68%) | 6 (75%) | 13 (65%) |  |
| ICI combination | 9 (32%) | 2 (25%) | 7 (35%) |  |
| Treatment Line (%) |  |  |  | 0.4 |
| 1 | 21 (75%) | 6 (75%) | 15 (75%) |  |
| 2 | 4 (14%) | 2 (25%) | 2 (10%) |  |
| 3+ | 3 (11%) | 0 (0%) | 3 (15%) |  |
| Treatment History |  |  |  |  |
| Prior ICI | 7 (25%) | 2 (25%) | 5 (25%) | >0.9 |
| Outcomes |  |  |  |  |
| Progression | 20 (71%) | 1 (13%) | 19 (95%) | <0.001 |
| Death | 18 (64%) | 2 (25%) | 16 (80%) | 0.011 |
| irAE | 19 (68%) | 8 (100%) | 11 (55%) | 0.029 |

<sup>1</sup> Mean (SD); n (%)<sup>2</sup> Wilcoxon rank sum test; Fisher's exact test

### Identifying time-dependent bacterial species associated with progression

In order to identify bacterial species that might predict ICI-resistance at the time of treatment initiation, we used a recursive feature elimination (RFE) with random forest classifiers to identify the top features that could distinguish fecal samples at the time of progression. Pre-treatment fecal samples (Table S1, timepoint T0) were reserved as a test set since predication prior to treatment initiation would be a more clinically meaningful biomarker. All other longitudinal samples were used to train the model. Samples were labeled based on based on temporal proximity to progression: “progression-associated” for samples collected ≤ 60 days of disease progression and “progression-unrelated” if collected >60 days from progression. Training variables included non-ambiguous bacterial species (n=145, Table S6) and 3 clinical covariates from Fig 1A (treatment line, TMB, and smoking status, p values < 0.01). Of note, since we wanted to develop this model for prediction prior to treatment initiation, we excluded irAE data from the training model despite significance as a co-variate (Fig 1A), as the ability to predict which patients will develop irAEs is not currently feasible.

With these parameters, the RFE identified 44 optimal features, including 42 species, treatment line and TMB, that were able to distinguish between progression-associated and progression-unrelated samples (Table S6). Of these, the 42 bacterial species were further filtered on fold change (abs[log_2_ fold change] ≥ 1) across treatment groups, resulting in a final list of 20 progression-associated species (Table S7). To validate the RFE model, a composite index of the 20 species was calculated for each sample as the ratio of cumulative abundances of "favorable" (log₂ fold change ≥ 1) versus "unfavorable" (log₂ fold change ≤ –1) features (Table S8). A ratio of “favorable” to “unfavorable” was applied to the test set of pre-treatment samples (n=50 samples for 50 individuals, Table S1, timepoint T0) and stratified based on median value. Patients with index score ≥ median had better PFS and OS, while patients with scores < median had worse PFS and OS (Fig 4A and B, respectively). We further validated this index using an independent cohort of NSCLC patients with pre-treatment samples (Fig 4C-D, n=70 patients)^19^, which was consistent with the results in our own cohort (Fig 4A-B, Table 1).

**Figure 4.**
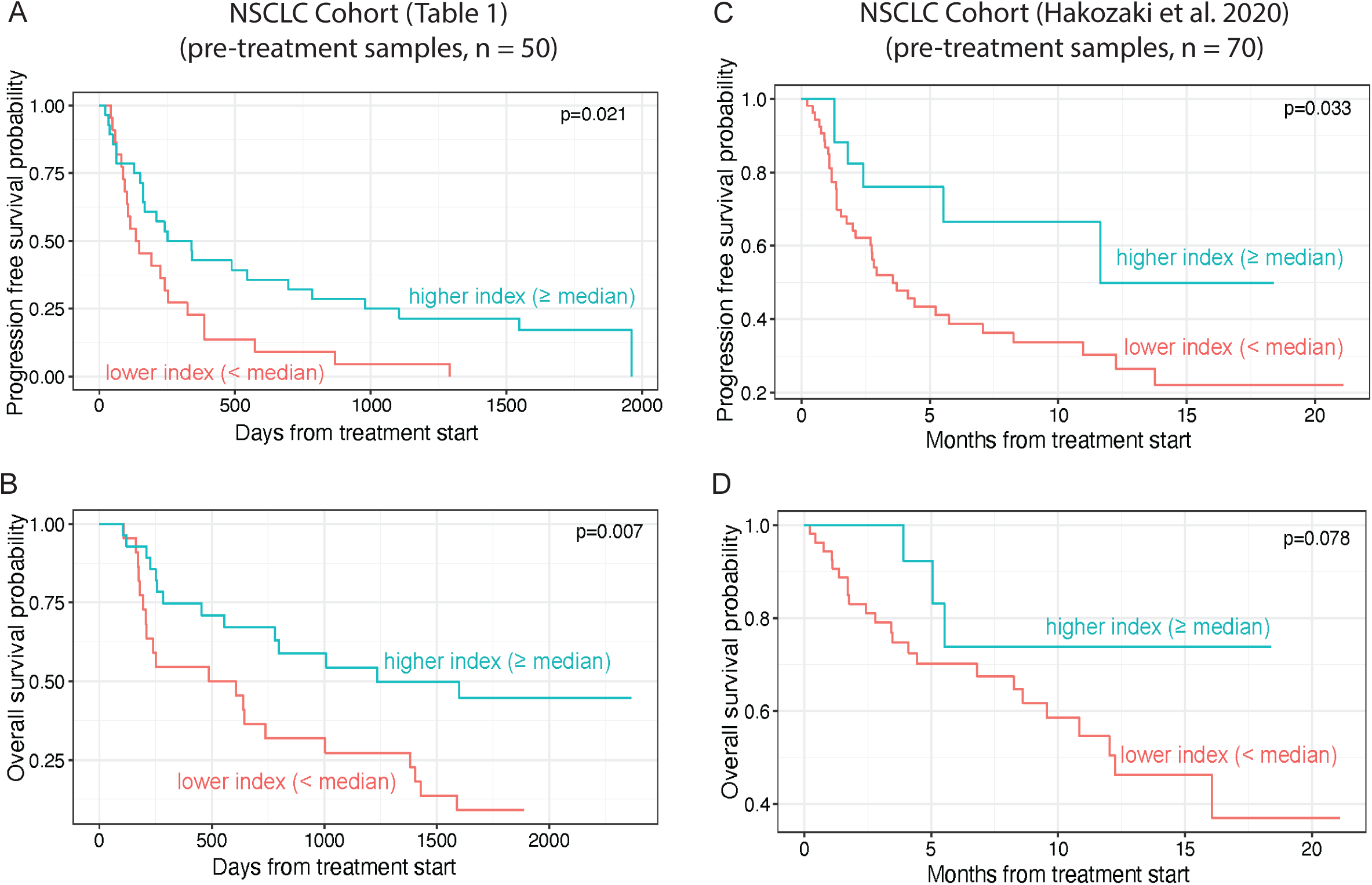
A progression-associated index of bacterial species is predictive of clinical outcomes. Microbiome and clinical covariate features from longitudinal samples (pre-treatment excluded) were ranked using a recursive feature elimination with random forest classifiers, which identified the 44 features (42 bacterial species, treatment line, tumor mutation burden) able to distinguish between progression-associated and - unrelated longitudinal samples. The 42 bacterial species (Table S6) were further filtered using relative abundance, and those with abs[log_2_ fold change] ≥ 1 were included in the final index of 20 species (Table S8). ***A-B.*** Kaplan–Meier probability curves for progression free survival (A) and overall survival (B) at pre-treatment according to high (≥ median) and low (< median) values of the microbiome index applied to fecal samples (n=50 patients). ***F-G*.** Kaplan–Meier probability curves for progression free survival (C) and overall survival (D) according to high (≥ median) and low (< median) values of the microbiome index applied to fecal samples. (n=70 patients)

## DISCUSSION

A key current obstacle to improving outcomes for patients with locally advanced or metastatic NSCLC is predicting and overcoming resistance to ICI-based regimens. The microbiome has been shown to be a potentially modifiable biomarker of tumor response, but the ability to apply such biomarkers has been limited by a lack of longitudinal data, as well as limited integration of other relevant clinical features that can affect both the microbiome itself (e.g. antibiotic use) and clinical outcomes from ICI-treatment in NSCLC (TMB, PD-L1). Our data begins to explore the longitudinal microbiome stability, correlating the microbiome alterations with treatment modality, treatment line, TMB, irAEs, and antibiotic exposure. In this study, we identify that fecal microbiome instability occurs in patients who experience primary resistance to ICI-based regimens within the first 4 months on therapy (120 days), particularly notable in patients on chemotherapy and in contrast to the microbiome changes that occur in patients who experience either acquired resistance or durable response while on treatment. Specifically, the microbiome in primary resistance changes rapidly within an individual, in both NSCLC and melanoma, and a lack of stability is associated with poor tumor response across both tumor types. Finally, we used microbiome species temporally associated with disease progression to develop a potential biomarker that may be predictive of early resistance to ICI-based regimens when applied prior to treatment initiation.

To date, the majority of microbiome cohort studies have utilized cross-sectional analysis at pre-treatment timepoints, though longitudinal data has been reported in two melanoma cohorts. Our results for microbiome stability are consistent with data reported for in melanoma, where patients who experience a complete response or PFS > 12 months were found to have a more stable microbiome^26,27^. The data for specific bacterial species, however, remains disparate between our study and other longitudinal cohorts. These datasets are further confounded by different tumor types and environmental factors, such as geographic location, which can contribute to diversity among bacterial species^34^. For example, in our NSCLC cohort, a longitudinal change in abundance of *Anaerostipes caccae* was associated with progression, while *A. caccae* was found to remain stably enriched in patients with melanoma who had a complete response. Other reports note stability in multiple Clostridia species associated with complete response, while our study indicates longitudinal shifts in specific Clostridia species (*C. clostridioforme and C. ruminantium)* associated with nonresponse. In contrast, however, both prior reports and our data suggest some concordant results using functional pathway analysis. Multiple menaquinol (vitamin K) pathways were found to be enriched in melanoma nonresponders and our NSCLC cohort^26^. Manequinol pathways have been associated with chronic inflammatory and cardiovascular diseases, perhaps suggesting impaired immune responses during treatment with ICI-based therapy^35^. This concordance across predicated functional pathways suggests that analysis of the aggregate bacterial community, rather than individual species, may yield more consistent results across cohorts.

Overall, these data suggest a potential opportunity to use the gut microbiome to identify patients with ICI-resistance early in their treatment course, at a timepoint when standard of care imaging may still be ambiguous or demonstrate unusual immune-related tumor dynamics, such as pseudo-progression^36^. To date, established biomarkers (e.g. PD-L1, TMB) have been useful in identifying patients with NSCLC who may benefit from ICI-based therapy^37^. However, these biomarkers do not distinguish ICI-resistance and are further limited in their ability to be repeatedly or longitudinally addressed due to the need for invasive biopsies. In contrast, sampling the gut microbiome is non-invasive, thus making longitudinal assessment more accessible during treatment. Identifying patients at risk for poor tumor response at an early timepoint could offer additional opportunities to develop microbiome-based interventions that aim to overcome ICI resistance, which remains the primary barrier to progress for ICI therapy in NSCLC.

Our study was also able to incorporate antibiotics exposure with fecal microbiome shifts, and interestingly, the longitudinal shifts in microbiome stability observed in those with primary resistance were not driven by antibiotic exposure. In fact, antibiotic exposure near the start of treatment start was not associated with worse PFS in our cohort, contrary to other published studies^30^. However, a subset analysis does suggest that antibiotic exposure may be more important in acquired resistance, compared to primary resistance. Several factors could account for these discrepancies. First the tumor biology in primary versus acquired resistance is likely related to multiple tumor-dependent factors, including the expression of multiple checkpoint inhibitors and a potentially altered tumor microenvironment^8^. Consistent with this idea, tumor-dependent factors may be a stronger driver of the immune response in primary resistance, whereas a combination of multiple factors, including antibiotic exposure, could impact ICI resistance at later timepoints for acquired resistance. Second, our cohort, like many previously published cohorts examining the effect of antibiotic exposure, remains a single-center analysis with samples collected at timepoints during which patients attended for therapy or standard of care assessments, rather than a dedicated prospective study. Prior studies have not typically examined different patterns of ICI resistance, and thus, further examination with pre-defined timepoints for specimen collection and a more robust investigation of antibiotic type, mode of administration and drug pharmacokinetics may be necessary to elucidate the nuances of antibiotic exposure on an ICI-based regimen.

When examining patterns of ICI resistance, our study was able to distinguish between primary and acquired resistance in NSCLC. However, our data for durable response is more limited, which may be due to smaller sample size in this group or longer duration of time over which samples were collected since patients with durable responses remained on therapy for longer than those with ICI resistance. Additionally, as above, the underlying biology of ICI response in patients with durable response and the clinical covariates that drive durable response may be distinct from primary or acquired resistance. For example, almost half of our durable response cohort had high TMB, which was associated with longer PFS (Table 1, Fig 1A). A dedicated multi-center study of durable response in NSCLC could further elucidate key factors driving long-term ICI responses.

In summary, these data indicate that longitudinal analysis and microbiome trajectory early in treatment with an ICI-based regimen may be a predictor of ICI resistance, especially as combination chemo-immunotherapy has become first line treatment in NSCLC without a driver mutation. Additional analysis with dedicated cohort studies across cancer types in the first line setting and pre-determined collection timepoints will be necessary to develop and validate a longitudinal microbiome-based predictive biomarker.

## METHODS

### Study design, eligibility criteria, and participants

All participants were enrolled at Johns Hopkins Hospital, Johns Hopkins Bayview Medical Center, Johns Hopkins Sibley Memorial Hospital, or Johns Hopkins Suburban Hospital (IRB00108798, IRB00112429). Eligibility criteria included age ≥ 18 years, pathologically- or clinically confirmed malignancy, treatment with an ICI-based regimen (anti-PD-1, anti-PD-L1, anti-CTLA-4), and willingness to provide biospecimens. Exclusion criteria included active bacterial or viral infection at the time of screening, active tuberculosis or patients on medication for tuberculosis, pregnancy, and inability to provide biospecimens. All study protocols and all amendments were approved by the Institutional Review Board of Johns Hopkins University. Written informed consents were obtained from all participants.

Patients with thoracic malignancies were enrolled between December 13, 2016, and July 14, 2023, with data lock on February 23, 2024. Patients with melanoma / skin malignancies were enrolled between September 28, 2017, and January 14, 2020, with data lock on May 9, 2024. Patients were selected for inclusion in this study based on a pathologically confirmed diagnosis of NSCLC or melanoma, respectively, with advanced/unresectable or metastatic disease. Patients undergoing active treatment for a second malignancy were excluded. Patients on both standard of care regimens and clinical trials were included. For the NSCLC cohort, a total of 112 patients with advanced/unresectable or metastatic NSCLC were identified, but 11 patients were excluded due insufficient sequencing reads from fecal samples (< 5000 cleaned paired-end reads after data processing) and/or inability to assess tumor response based on imaging studies. For the melanoma cohort, a total of 154 patients were identified who received systemic therapy for advanced/metastatic melanoma and/or unresectable disease. Of these, 28 patients had available pre-treatment and longitudinal fecal samples within 180 days of treatment start, allowing for direct comparison of microbiome profiles within each patient.

Cohort characteristics were extracted manually from the electronic medical records and/or REDCap databases. Tumor response was clinician-determined based on imaging studies, with ambiguous responses resolved by two or more medical oncologists. For the NSCLC cohort, tumor response was categorized into 4 groups based on PFS: primary resistance to an ICI-based regimen (n=47, PFS < 6 months), acquired resistance after 6 months of partial response or stable disease (n=19, PFS > 6 months but < 12 months), acquired resistance after 12 months of partial response or stable disease (n=21, PFS > 12 months with eventual progression), and durable response (n=14, PFS >12 months without progression at time data was censored). For the melanoma cohort, tumor response was categorized into responders or nonresponders based on response at 6 months.

### Biospecimen collection, DNA extraction, and sequencing

All fecal samples were collected by patients at home as either fresh stool or placed into an OMNIgene GUT tube (DNA Genotek, Stittsville, ON, USA, OM-200) at the time of collection. Fresh stool was stored at 4°C in a specimen cup and aliquoted in a laminar flow hood and then frozen at -80 °C within 48 hours of collection. OMNIgene GUT tubes were mailed or brought to the lab within 60 days, vortexed, and aliquoted in an anaerobic hood. Pre-treatment samples were defined as collected on day 1 of treatment initiation or up to 14 days prior to treatment initiation. For patients with multiple pre-treatment samples in this time range, the sample collected closed to treatment start date was defined as the pre-treatment sample. For longitudinal analysis, the inclusion criteria required at least 2 fecal samples available for analysis at -14 to 180 days, relative to treatment start. Patients with at least one pre-treatment sample were included for all analyses using pre-treatment samples only and for survival analysis.

For genomic DNA extraction, approximately 80mg of fresh stool or 250uL of OMNIgene GUT stool sample were transferred to BashingBead Lysis Tubes (Zymo, 0.1/0.5 mm beads) with bead bashing buffer, followed by mechanical lysis using a Mini-Beadbeater-96 (Biospec Products, Bartlesville, OK, USA) at 2400 rpm for 60 seconds, 3 cycles in total. The remainder of the DNA extraction was performed using the Quick-DNA Fecal/Soil Microbe Kit (Zymo, Irvine, CA, USA, 96 well plate). All sequencing was performed by the CHOP Microbiome Center as previously described^38^. All samples were stored at −80 °C. Sample and patient details are shown in Table S1.

### Sequencing data processing, analysis, and statistics

High-quality 16S rRNA amplicon sequences were assigned to a high-resolution taxonomic lineage using Resphera Insight (v.2.2), for which the performance has been previously benchmarked and described in detail^16,39^. Briefly, this method utilizes a manually curated 16S rRNA database of 11,000 unique species, and a hybrid global-local alignment procedure to assign short next-generation sequencing sequences from any region of the 16S rRNA gene to a high-resolution taxonomic lineage.

Sequences that are too divergent to be confidently classified to any species are subsequently clustered *de novo* into novel operational taxonomic units (OTU) using a 97% identity threshold and assigned a higher-level taxonomy. In this study, ambiguous assignments and novel OTU assignments were used for calculation of percentage abundance estimates per sample but were not considered as candidates for index discovery process. Ambiguous OTUs were included in all analyses and are shown in supplementary tables; however, ambiguous OTUs were excluded from final visualizations (Figures 1D, S9).

Alpha diversity measures were calculated using QIIME (v.1.8.0) and R package phyloseq (v1.42.0). Beta diversity was calculated based on Bray-Curtis, weighted UniFrac, and unweighted UniFrac distance using R package Vegan (v2.6.4). Functional profiles were predicted using PICRUSt2 (v.1.0.0) based on Metacyc databases^28^. Statistical analyses were performed using R (v4.2.2). The Mann–Whitney test was used to identify significant bacterial taxa associated with response groups at each timepoint and corrected for false discovery rate (FDR, adjusted p value < 0.05). Significant taxa from pairwise comparisons were reviewed based on prior occurrence in human studies based in literature search using www.pubmed.gov. Principal coordinates analysis (PCoA) was conducted using the ade4 package (v1.7-22) to visualize variance in bacterial taxa. Permutational multivariate analysis of variance (PERMANOVA) was performed using the vegan package (v2.6-4). Comparisons across > 2 groups were performed using a Kruskal–Wallis test. All visualizations were generated using the ggplot2 (v3.4.4), ggpubr (v0.6.0), and swimplot (v1.2.0) packages. Quantitative PCR for *Escherichia coli* was performed as previously described^40^. Taxonomic and pathway tables for all samples are available at https://github.com/fshaikh14/Shaikh-NSCLC-ICI-resistance.

### Antibiotic data extraction

Medication data was extracted from electronic medical records through The Lung Cancer Precision Medicine Analytics Platform (PMAP), which is a cloud-based data integration and analytics environment developed in the Lung Cancer Precision Medicine Center of Excellence (PMCOE) at Johns Hopkins University School of Medicine. Demographic, clinical, pathological, genomic, radiological, treatment, and outcome data from the electronic health record into an integrated, centralized data lake for transformation, projection, and analysis in secure workspace. All activities are conducted under the Lung Cancer PMCOE IRB approval (IRB00303251).

### Clinical co-variate and antibiotic analysis

To assess whether clinical covariates or antibiotic usage were associated with PFS, we applied univariate Cox proportional hazards regression models using the coxph() function from the R package survival (v3.7). Time-to-event data were defined from the start of therapy to either (1) progression or death from any cause (PFS) or (2) death from any cause (OS). We used these models to estimate hazard ratios (HRs) with 95% confidence intervals (CI) for OS. Survival curves were estimated using the Kaplan–Meier method via the survfit2() function from the R package ggsurvfit (v1.1.0). Antibiotic exposure was based on any antibiotic use within 42 days before or after treatment initiation, with further stratification based on route of administration (oral or intravenous/intramuscular). Antibiotic exposure near the time of progression was based on any antibiotic usage within 42 days prior to disease progression.

### Microbiome stability analyses

To evaluate the stability of bacterial composition within each individual across different response groups, intraclass correlation coefficient (ICC) was implemented using the ICC() function from the psych package (v2.4.6.26)^41^. Only patients with more than 2 samples were included in the analysis to ensure meaningful within-individual comparisons. To assess temporal shifts in the gut microbiome composition within individuals, we performed pairwise principal coordinate analysis (PCoA) using Bray-Curtis dissimilarity as above. For both primary resistance and acquired resistance group, we subset patient samples at specific timepoints (e.g., T0 vs. T60 or T0 vs. T120). For direct intra-individual comparisons, only individuals with paired samples across both time points were included in the analysis to ensure within-individual comparisons and the genus-level abundances were averaged if patients had multiple samples within a timepoint. Statistical significance of compositional change over time was tested using PERMANOVA (same as above) with timepoint as the main effect and record ID as the covariate. An additional metric of self-similarity was added as an aggregate measure over time of the shared taxa from pre-treatment, with change represented as the average of the relative abundance of the taxa that are shared between samples from the same patient at each timepoint. Individual data points were fit into locally estimated scatterplot smoothing (LOESS) with 95% confidence intervals, and bootstrapping (10 iterations to avoid overfitting dataset) was applied at days 30, 60, 90, and 120, followed by statistical testing using Mann–Whitney test (R base stats package, v4.1.1).

### Recursive Feature Elimination and progression-associated microbiome index

Recursive feature elimination (RFE) was applied to identify key bacterial species and clinical factors associated with progression status. The training set included only on-treatment samples (all samples that were not pre-treatment, i.e. samples not at T0, n = 324). Fecal samples collected ≤ 60 days of progression were defined as “progression-associated” while fecal samples collected >60 days from progression were “progression-unrelated”. Operational taxonomic units (OTUs) were aggregated by distinct species, and ambiguous assignments were removed prior to model training. Microbiome data were filtered to retain species with a relative abundance >0.01 and present in at least 10% of samples. Missing values in the numeric clinical variable (treatment lines) were imputed using the median and flagged with a binary indicator, while missing values in categorical variables (tumor mutation burden and smoking history) were replaced with “unknown” to retain training samples. RFE with randomForest classifiers was performed using the rfe() function from the caret package (v6.0-94), with repeated five-fold cross-validation (five repeats) for model evaluation. To identify when model accuracy plateaus with increasing number of features, we fit a smoothing spline to the accuracy results from RFE. The elbow (plateau) point was defined as the first index at which the second derivative changed sign. The model identified 44 optimal variables (42 species + treatment line and tumor mutation burden) based on cross-validation accuracy. The 42 species features were further filtered by requiring a mean absolute log₂ fold change ≥1 across response groups, based on pairwise Mann–Whitney tests, which results in 20 species. For validation, a composite index of 20 species was calculated for each sample as the ratio of cumulative relative abundances of "favorable" (log₂ fold change ≥ 1) versus "unfavorable" (log₂ fold change ≤ –1) features. To test, T0 samples (n=50 samples for 50 individuals, Table S1) were stratified into "higher" and "lower" index groups using the median index value from the training set as the cutoff. Survival analysis was conducted as described above. To validate in an independent cohort, the prognostic value of these 20 species was applied to an independent published dataset^19^ with data processing as previously described^16^.

## Data Availability

Authors consent to sharing of data and materials upon reasonable request to the authors.

## Declarations

### Ethics approval and consent to participate

All studies were approved by Institutional Review Boards at Johns Hopkins University School of Medicine, sites include Johns Hopkins Hospital, Johns Hopkins Bayview Medical Center, Johns Hopkins Sibley Memorial Hospital, and Johns Hopkins Suburban Hospital (IRB00108798, IRB00112429). All identifiers in this manuscript (SampleID, RecordID) were not known to anyone outside the research team. The sample and patient identifiers were assigned independent of patient medical records and only used for the purposes of this research study. All timepoints are provided as relative dates to treatment start.

### Consent for publication

All authors have approved the following manuscript prior to submission

### Availability of data and material

Authors consent to sharing of data and materials upon reasonable request to the authors.

### Competing interests

F.Y.S. has received research funding to Johns Hopkins University from Bristol-Myers Squibb. J.R.W. reports equity ownership of Resphera Biosciences. E.J.S. is a consultant for Boston Scientific. D.M.P. reports stock/ownership in Aduro Biotech, Dracen, Ervaxx, Five Prime Therapeutics, Tizona, Trieza Therapeutics, and WindMil. C.L.S has received research funding to Johns Hopkins University from Bristol-Myers Squibb, and royalties from Up to Date outside the submitted work. JN has received research funding from AstraZeneca Bristol-Myers Squibb, Roche/Genentech, Takeda, Pfizer, Novartis, Mirati, Summit Therapeutics; consulting/advisory boards from: AstraZeneca Bristol-Myers Squibb, Roche/Genentech, Merck, Mirati, Regeneron, Daiichi Sankyo, Zymeworks, Summit Therapeutics; honoraria from AstraZeneca Bristol-Myers Squibb, Roche/Genentech, Merck, Mirati, Regeneron, Daiichi Sankyo, Zymeworks, Summit Therapeutics, and Data Safety Monitoring Boards/Steering Committees for: AstraZeneca Bristol-Myers Squibb, Daiichi Sankyo, Summit Therapeutics.

### Funding

The translational work in this manuscript and authors were supported by the following: Bloomberg∼Kimmel Institute for Cancer Immunotherapy (D.M.P, C.L.S, F.Y.S.), a research grant from Bristol Myers Squibb (D.M.P., C.L.S.), US National Institutes of Health (K08CA263316 to F.Y.S), Damon Runyon Clinical Investigator Award (F.Y.S), The Marilyn and Michael Glosserman Fund for Basal Cell Carcinoma and Melanoma Research (E.J.L., M.D.S.), The Barney Family Foundation (E.J.L., M.D.S., W.H.S.), The Laverna Hahn Charitable Trust (E.J.L., M.D.S., W.H.S.), and an IASLC Lung Cancer Foundation of America Young Investigator Award (JN).

### Authors’ contributions

*C.L.S, D.M.P, J.N., E.J.L.* conceived of the conceptual framework and contributed to the original study design. *J.G., S.G., W.O.A., K.P., M.S., W.H.S., P.F., K.A.M., E.J.L., J.N., and C.L.S*. were involved in the clinical study, enrollment, sample processing. *F.Y.S., M.C., J.T.F., J.N, E.J.L., C.L.S, Y.Z., J.R.W., K.C., J.M*. were directly involved in data collection, study design, analysis. *F.Y.S.* and *Y.Z.* wrote the first draft of the manuscript. All authors contributed to data interpretation, editing of the manuscript, and approved the final version of this manuscript.

## Acknowledgements

The authors wish to thank S. Rice, M. Leak, R. Carlson, and their respective administrative teams, and C. Barkley and I. Beadles for assistance for sample collection.

## LIST OF ABBREVIATIONS

ICI: immune checkpoint inhibitor
NSCLC: non-small cell lung cancer
STK11: Serine/threonine kinase 11
PFS: progression free survival
OS: overall survival
PD-1: programmed death 1
PD-L1: programmed death ligand 1
CTLA-4: cytotoxic T-lymphocyte-associated protein 4
OTU: operational taxonomic unit
FDR: false discovery rate
qPCR: quantitative polymerase chain reaction
PICRUSt2: Phylogenetic Investigation of Communities by Reconstruction of Unobserved States
HR: hazard ratio
CI: confidence interval
ICC: intraclass correlation
LOESS: locally estimated scatterplot smoothing
RFE: recursive feature elimination
CTLA-4: cytotoxic T-lymphocyte-associated protein 4
PR: primary ICI resistance
AR: acquired ICI resistance after an initial response of 6 months
AR6: acquired ICI resistance after 6 months of response
AR12: acquired ICI resistance after 12 months of response
ECOG: Eastern Group Cooperative Score
irAE: immune-related adverse event
TPS: tumor proportion score
TMB: tumor mutation burden
RFE: recursive feature elimination
PMAP: Lung Cancer Precision Medicine Analytics Platform
PMCOE: Precision Medicine Center of Excellence
T0: pre-treatment samples
T60: samples collected on days 1-60
T120: samples collected on days 61-120
T180: samples collected on days 121-180
T240: samples collected on days 181-240
T300: samples collected on days 241-300
T360: samples collected on days 301-360
T420: samples collected on days 361-420
T480: samples collected on days 421-480
T540: samples collected on days 481-540
T600: samples collected on days 541-600

---

**Figure S1.**
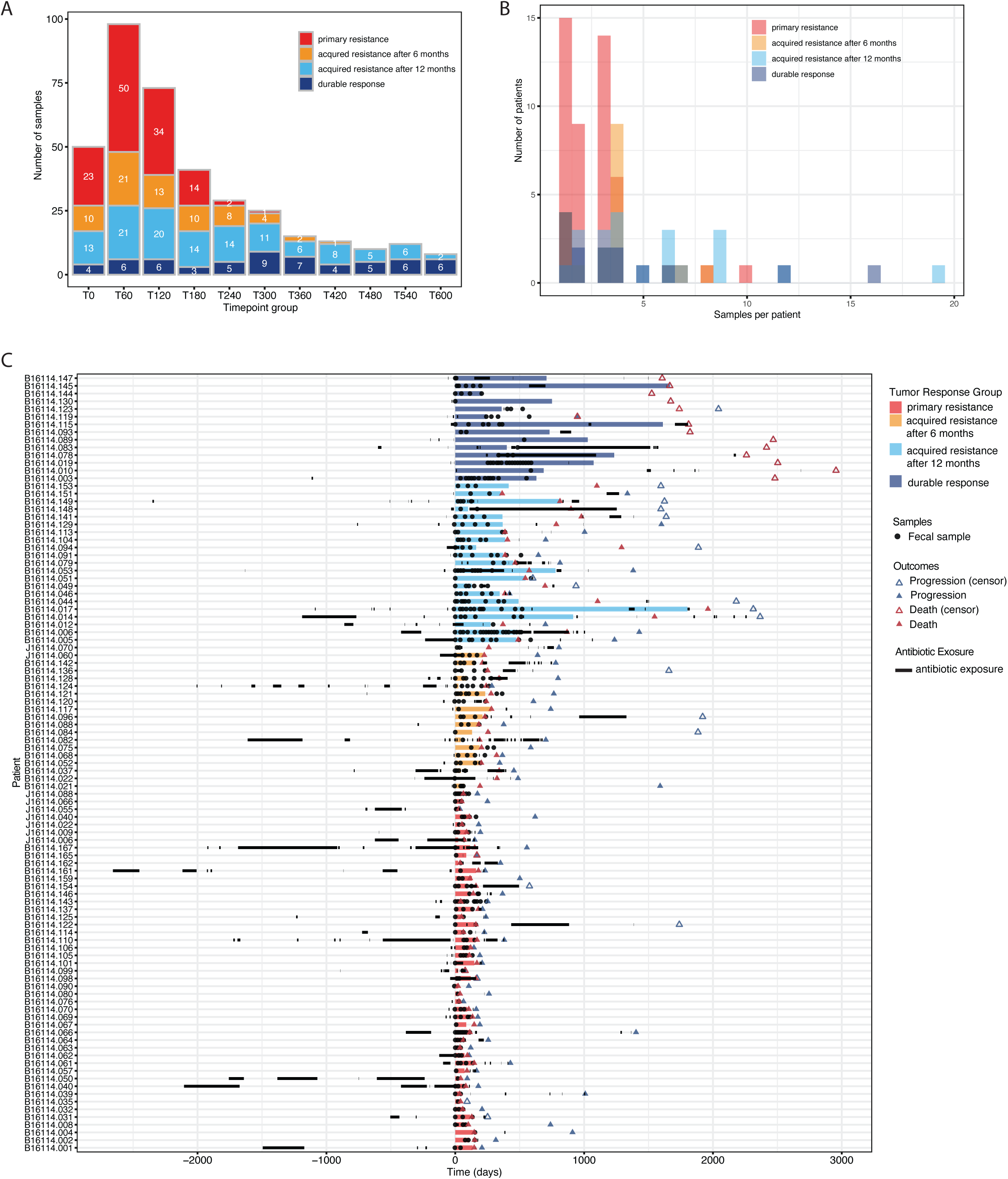
Distribution of samples for non-small cell lung cancer cohort. ***A***. Total samples by response group at each timepoint. ***B***. Total samples per patient by response group. ***C***. Samples per patients are shown relative to treatment start and annotated for progression, death, censor date, and antibiotic usage. ***Abbreviations***. PR, primary resistance; AR6, acquired resistance after 6 months of partial response or stable disease, AR12, acquired resistance after 12 months of partial response or stable disease; DR, durable response. ***Timepoints***. All timepoints are relative to treatment start date for each patient. T0, pre-treatment samples; T60, samples collected on days 1-60; T120, samples collected on days 61-120; T180, samples collected on days 121-180; T240, samples collected on days 181-240; T300, samples collected on days 241-300; T360, samples collected on days 301-360; T420, samples collected on days 361-420; T480, samples collected on days 421-480; T540, samples collected on days 481-540; T600, samples collected on days 541-600.

**Figure S2.**
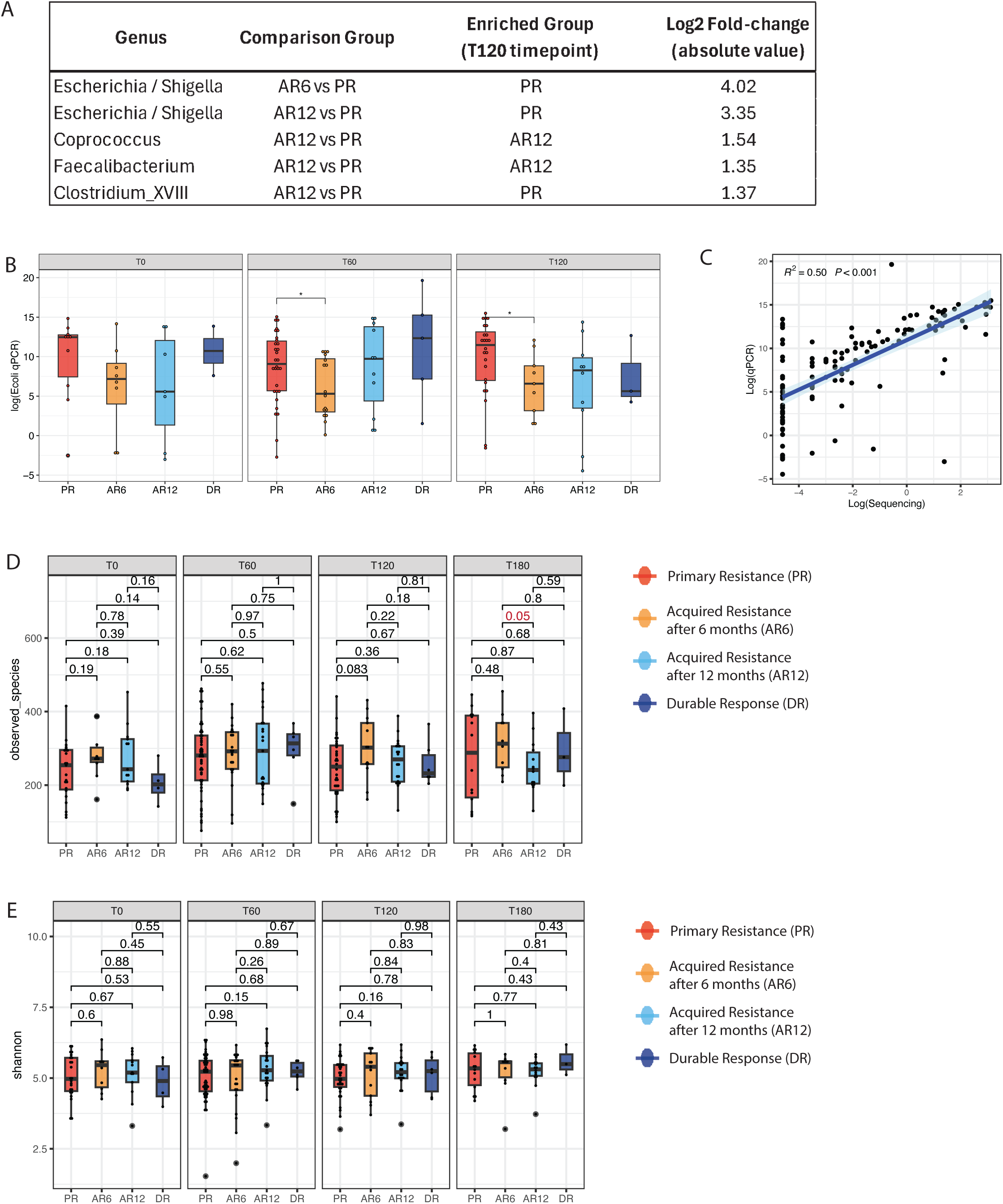
Microbiome features by treatment response group and timepoint. ***A.*** All unambiguous genera identified by Wilcoxon rank sum test atter correction for false discovery rate (p_adj_<0.05) in treatment groups at T120 (days 61-120). Table shows group comparison, enriched group, and absolute value of fold-change. Results correspond with Table S4. ***B***. Abundance of the genus *Escherichia* was quantified using quantitative PCR, shown at each timepoint (panel) by response group (color). Y axis indicates log-transformed colony forming units using a standard curve. Boxplot represents median, 25% to 75% interquartile range (bottom and top edge), and 95% confidence intervals (whiskers). **C**. Spearman correlation for genus Escherichia as detected by 16S rRNA amplicon sequencing and qPCR. ***D-E***. Alpha diversity across fecal samples by treatment response group (colors) and timepoints (panels). Observed Species (D) Shannon Index (E) are shown. Statistics: Wilcoxon Rank Sum.

**Figure S3.**
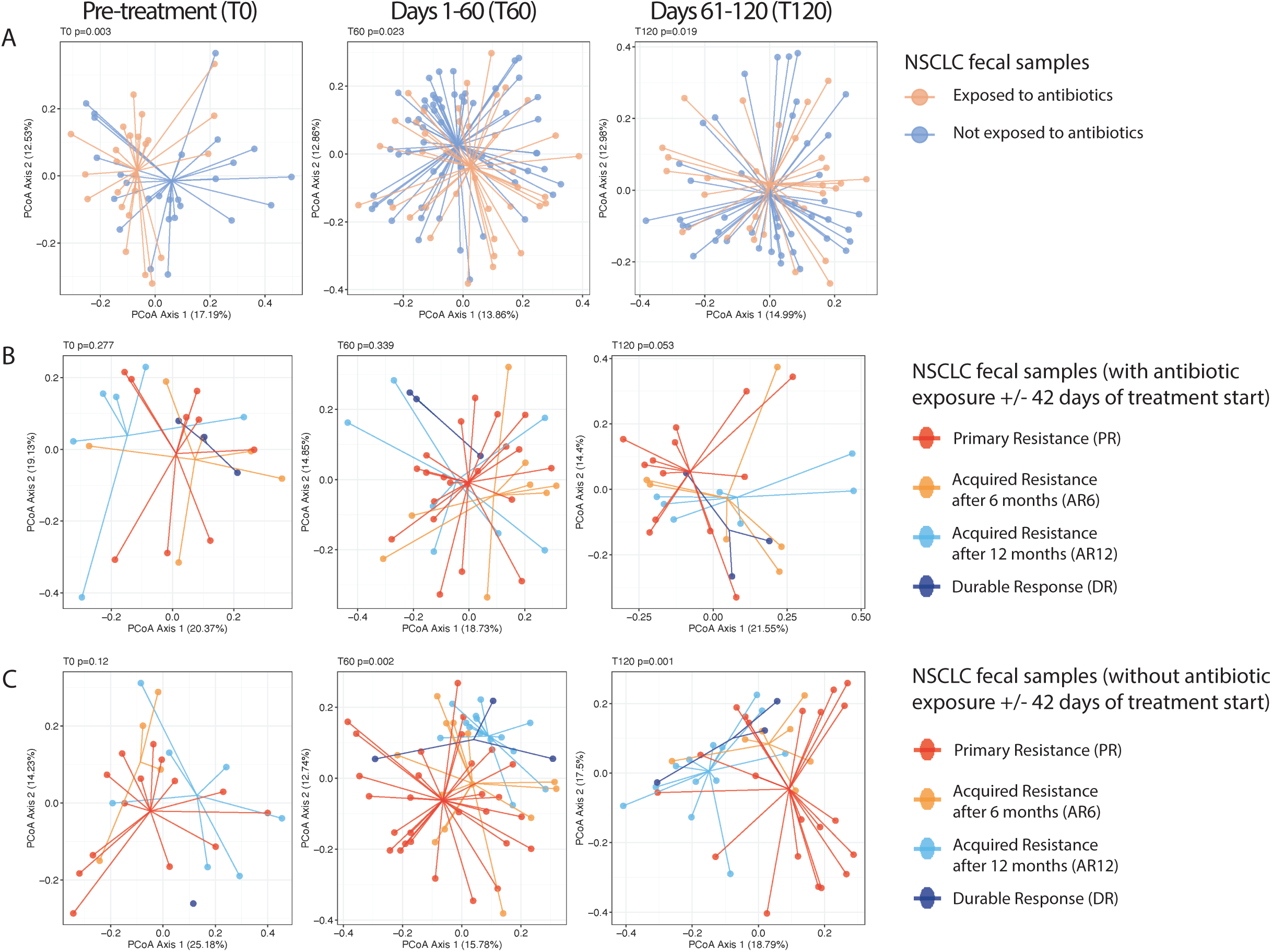
Microbiome β diversity stratified by antibiotic exposure +/- 42 days of treatment initiation. Principal Coordinate Analysis (PCoA) of fecal samples representing Bray-Curtis distance metric across timepoints. All timepoints are relative to treatment start date for each patient. T0, pre-treatment samples; T60, samples collected on days 1-60; T120, samples collected on days 61-120. Statistics comparison performed using Permutational Multivariate Analysis of Variance (PERMANOVA). ***A***. All samples stratified by patient antibiotic exposure. ***B***. All samples for patients exposed to antibiotics stratified by treatment response group. ***C***. All samples for patients not exposed to antibiotics stratified by treatment response group. Abbreviations. PR, primary resistance; AR6, acquired resistance after 6 months of partial response or stable disease, AR12, acquired resistance after 12 months of partial response or stable disease; DR, durable response.

**Figure S4.**
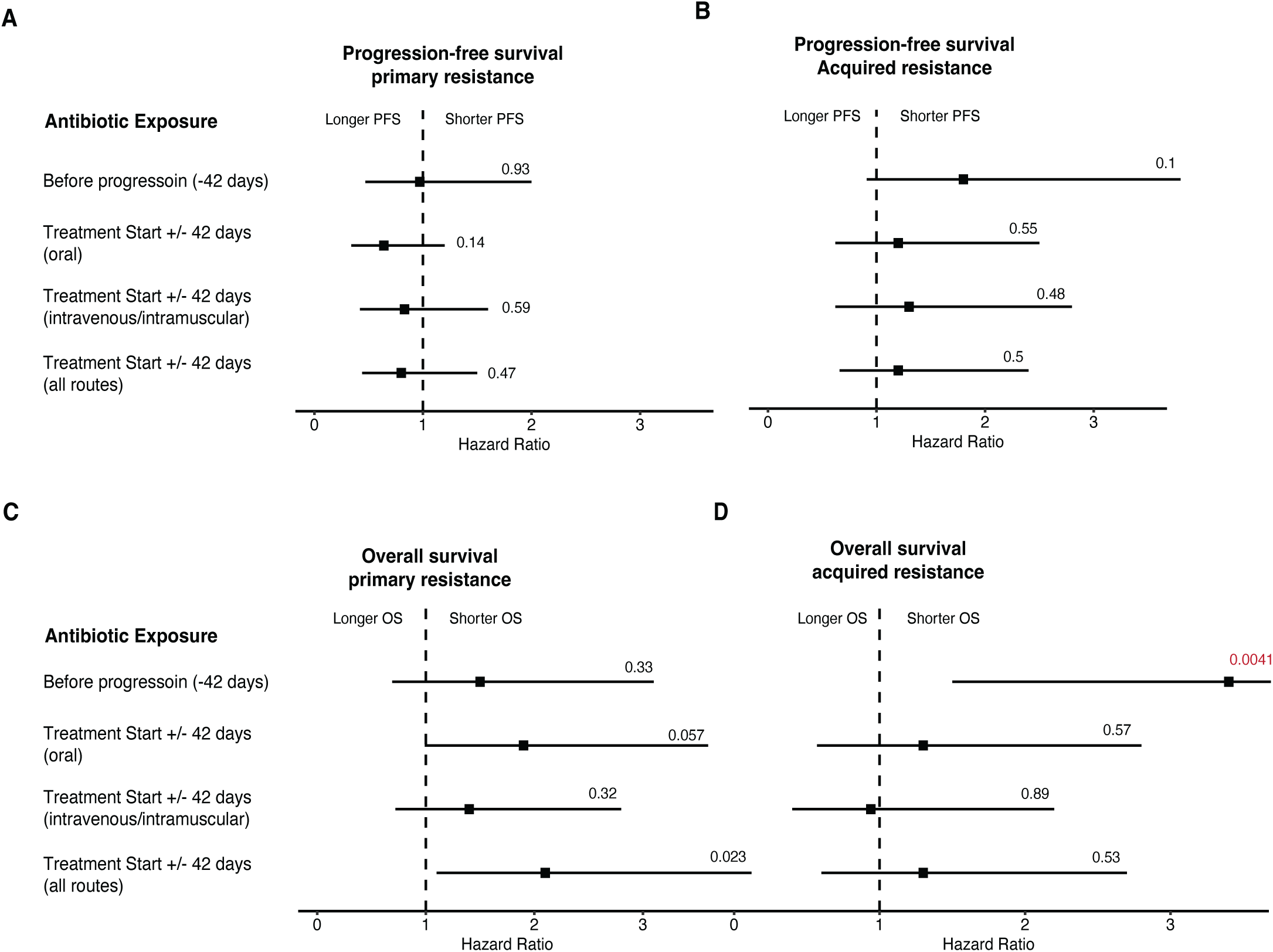
Antibiotic use within 42 days of progression was associated with worse clinical outcomes in acquired resistance. Cox regression analysis relative to progression free survival (PFS) and overall survival (OS) for primary resistance (A-B) and combined acquired resistance groups (C-D). Square represents the hazard ratio and the horizontal bars represent the 95% confidence interval. Statistics: likelihood ratio test.

**Figure S5.**
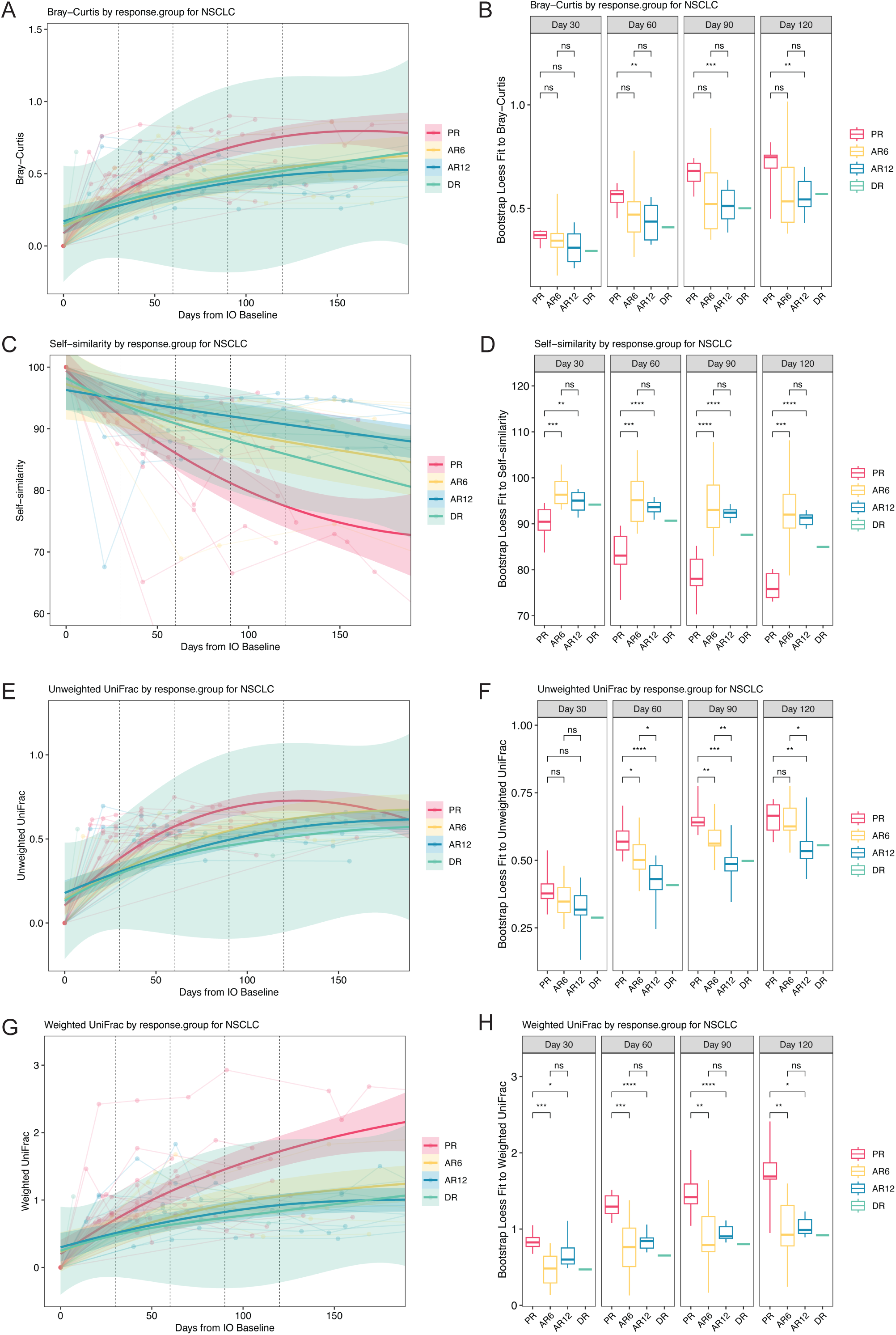
NSCLC primary resistance is associated with a loss of intra-individual taxonomic stability compared to acquired resistance. NSCLC fecal samples were assessed for intra-individual taxonomic stability using multiple similarity metrics. Distance metrics are shown using a locally estimated scatterplot smoothing (LOESS) model for fecal samples stratified by tumor response groups: primary resistance (PR), acquired resistance after 6 months of partial response or stable disease (AR6), acquired resistance after 12 months of partial response or stable disease (AR12), and durable response (DR). Individual samples are shown with lines connecting each patient. Data from LOESS curves was quantified at days 30, 60, 90, and 120 using bootstrap. ***A-B***, Bray-Curtis dissimilarity. ***C-D***, self-similarity. ***E-F***, unweighted UniFrac. ***G-H***, weighted UniFrac.

**Figure S6.**
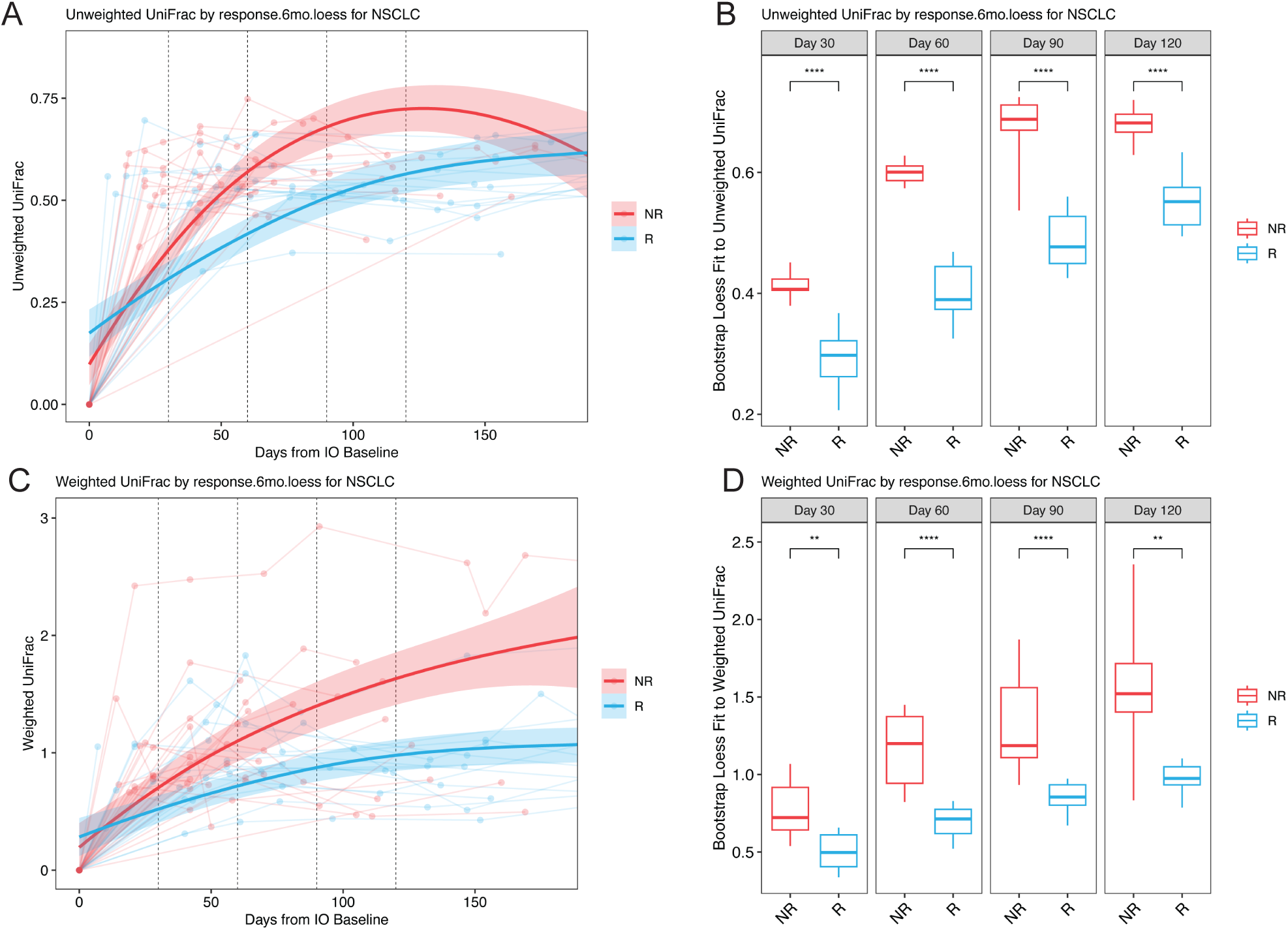
NSCLC nonresponse is associated with a loss of intra-individual taxonomic stability. Distance metrics are shown using a locally estimated scatterplot smoothing (LOESS) model for fecal samples sorted by tumor response at 6 months into nonresponders (primary resistance) and responders (acquired resistance and durable response). Individual samples are shown with lines connecting each patient. Data from LOESS curves was quantified at days 30, 60, 90, and 120 using bootstrap. ***A-B***, unweighted UniFrac distance, ***E-F***, weighted UniFrac distance.

**Figure S7.**
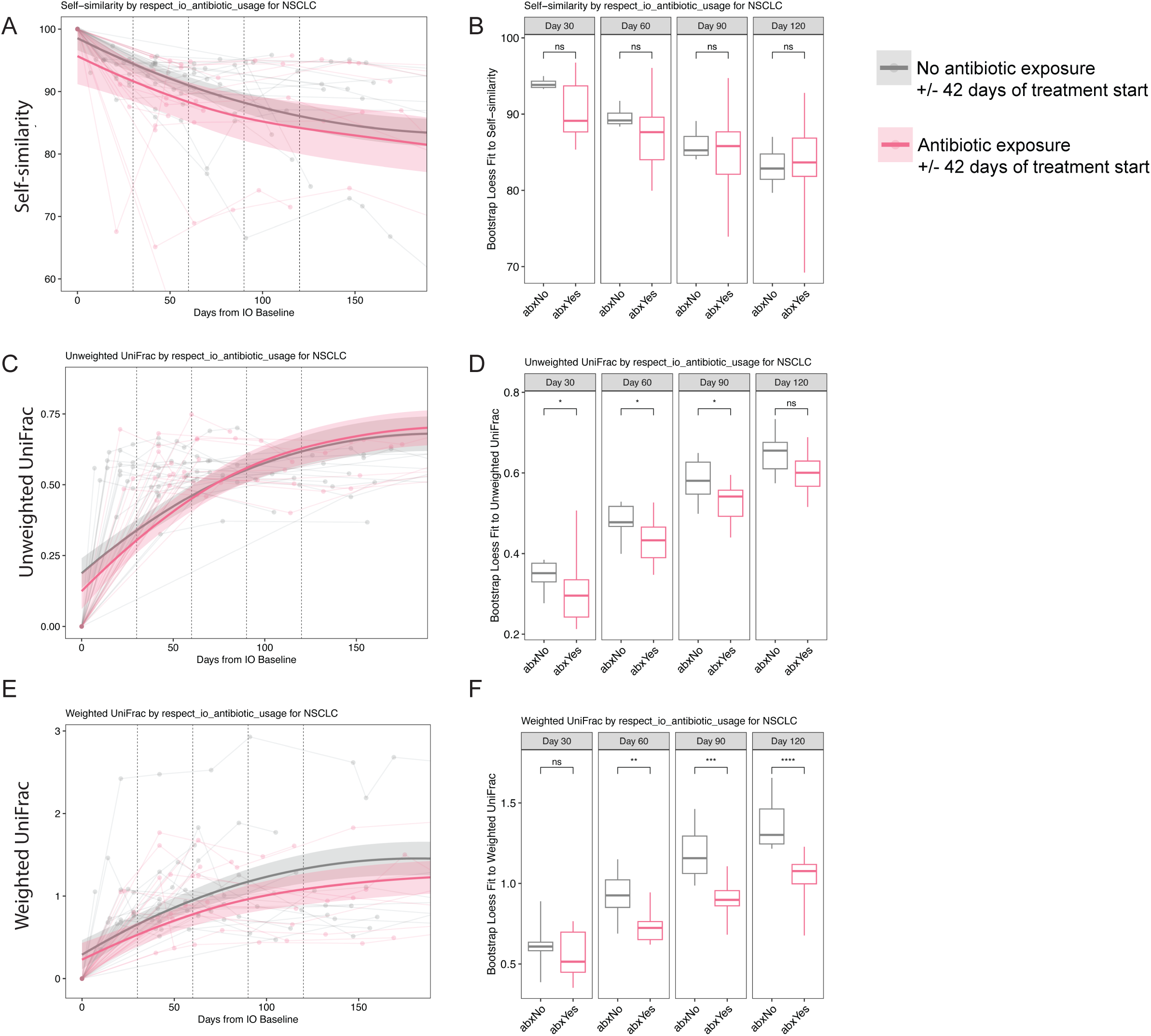
Antibiotic exposure does not drive intra-individual taxonomic instability. Distance metrics are shown using a locally estimated scatterplot smoothing (LOESS) model for fecal samples stratified by antibiotic exposure. Individual samples are shown with lines connecting each patient. Data from LOESS curves was quantified at days 30, 60, 90, and 120 using bootstrap. ***A-B***, self-similarity. ***C-D***, unweighted UniFrac distance, ***E-F***, weighted unifrac distance.

**Figure S8.**
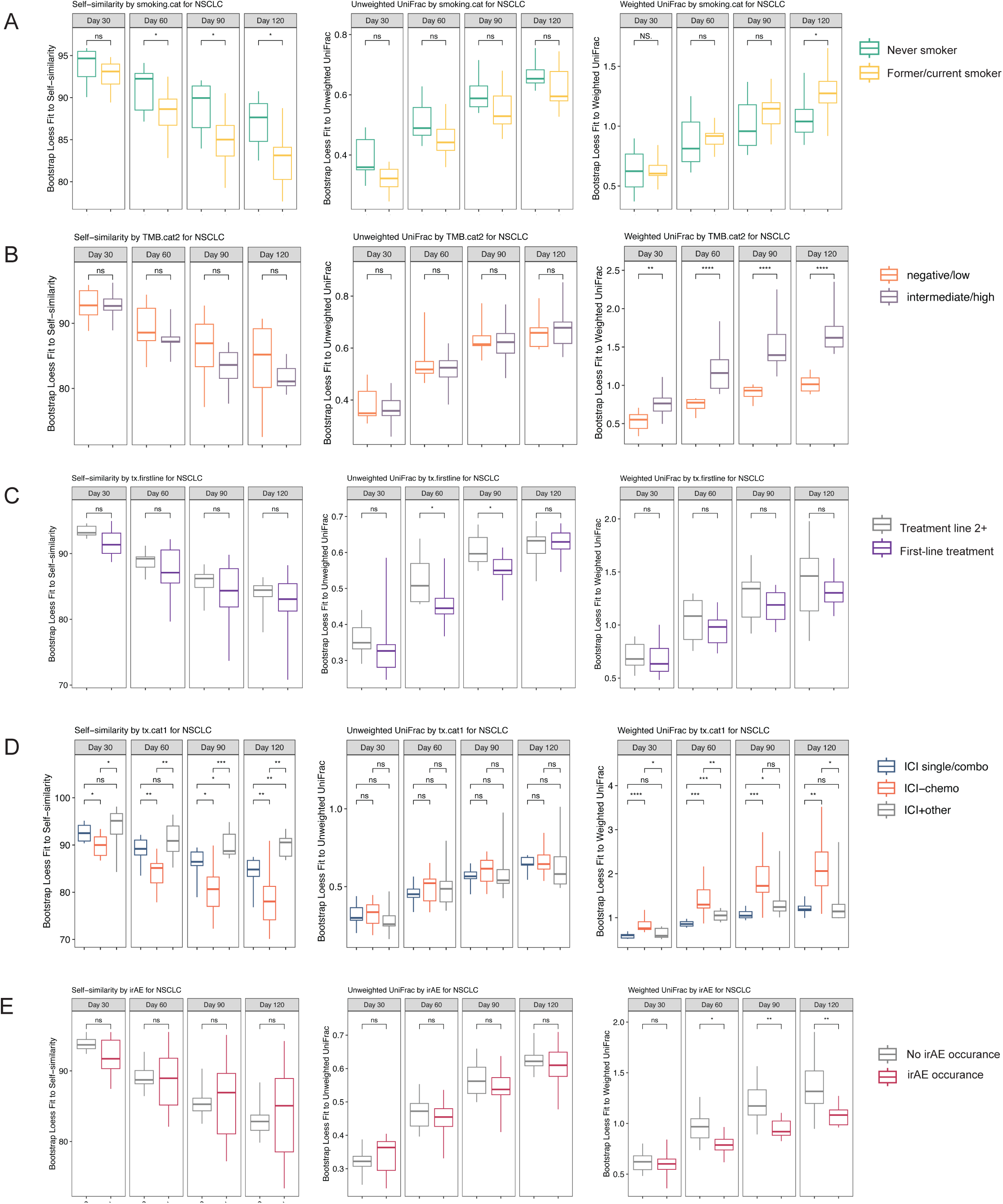
Clinical co-variates affect intra-individual taxonomic stability using multiple metrics. Distance metrics for fecal samples were fit to a locally estimated scatterplot smoothing (LOESS) model and stratified by the indicated clinical co-variate. Data shown is the quantification from LOESS curves at days 30, 60, 90, and 120 using bootstrap (left, self-similarity; middle, unweighted UniFrac; right, weighted UniFrac). ***A***, Smoking status. ***B***, tumor mutation burden (TMB). ***C***, treatment line. ***D***, treatment modality. ***E***, immune-related adverse events (irAE).

**Figure S9.**
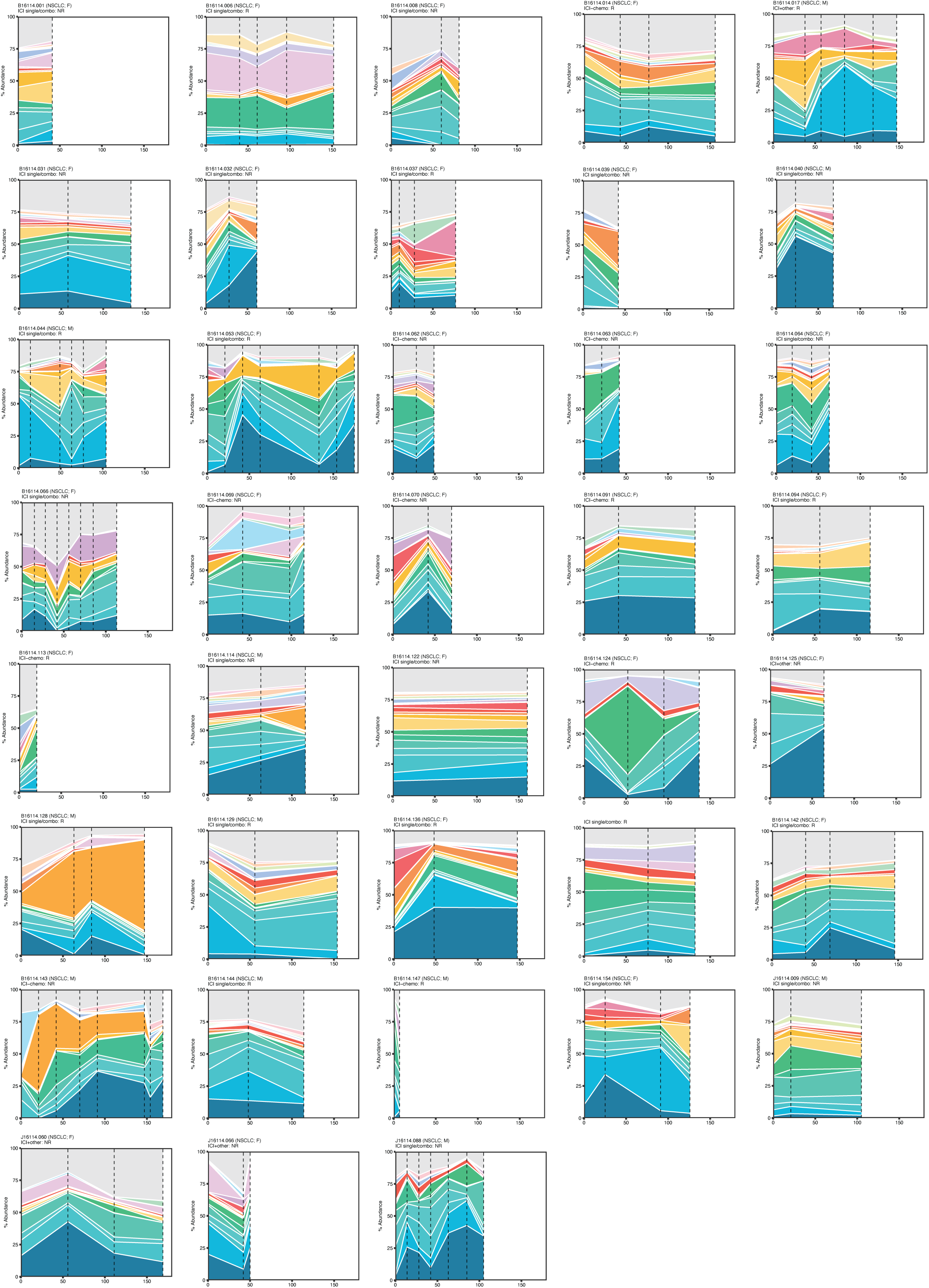
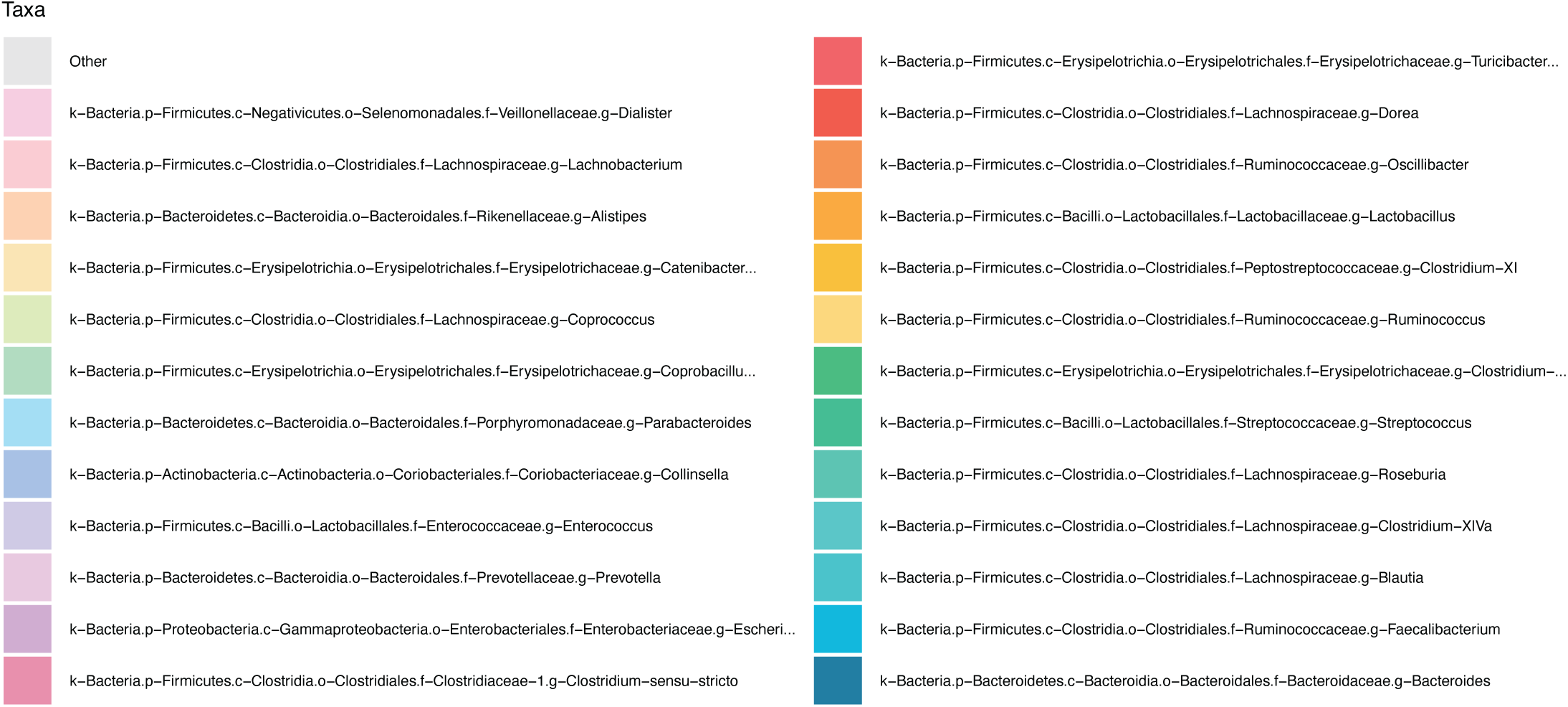
Dynamics of taxonomic classification shown longitudinally for patients with non-small cell lung cancer (NSCLC) who had available pre-treatment samples by genera (n=38). Timepoints indicated on x-axis, with all pre-treatment samples shown at day 0. Vertical axis is relative abundance by genus. Dotted vertical lines indicate fecal sample collected.

## SUPPLEMENTAL TABLES

**Table S1.** Sample manifest with associated clinical data.

**Table S2.** Cox proportional hazard relative to progression-free survival.

**Table S3.** Cox proportional hazard relative to overall survival.

**Table S4.** Significant genera at T120 (days 61-120)

**Table S5.** Longitudinal fold change (log2) enrichment of PICRUSt2 pathways in primary resistance

**Table S6.** Relative abundance of non-ambiguous species (n=145) for recursive feature elimination training model

**Table S7.** Importance scores using a random forest training model

**Table S8.** Fold change across all treatment groups for the 20 species used in progression-associated index

## Notes

### Author Declarations

The Institutional Review Boards at Johns Hopkins University School of Medicine (sites include Johns Hopkins Hospital, Johns Hopkins Bayview Medical Center, Johns Hopkins Sibley Memorial Hospital, and Johns Hopkins Suburban Hospital) gave ethical approval for this work.

